# Safety and efficacy of high frequency jet ventilation: a systematic and narrative review

**DOI:** 10.1101/2024.12.03.24318290

**Authors:** Jasmin Spaar, Peter Biro, Michael Sander, Volker Gross, Michael Scholtes, Keywan Sohrabi

## Abstract

High frequency jet ventilation (HFJV) is an unconventional mechanical ventilation technique increasingly utilized in complex medical fields such as airway surgery and intensive care.

This systematic review analyzes the safety and efficacy of HFJV, focusing on its application in both adult and pediatric populations. Through systematic searches of PubMed, Cochrane Library, and Livivo databases, 41 studies meeting the inclusion criteria were identified. Key parameters analyzed included complications, CO₂ and O₂ levels, pH values, FiO₂, treatment course, and duration. Studies were selected that evaluated HFJV both as a standalone method and in comparison, with conventional ventilation techniques.

The findings suggest that HFJV offers significant advantages in maintaining low alveolar pressures and improving surgical conditions, particularly in procedures requiring minimal organ movement. While most studies reported no significant difference in complication rates between HFJV and conventional ventilation, HFJV was associated with a more favorable treatment course and duration. Despite these positive outcomes, the data highlight challenges in the use of HFJV, particularly the complexity of the technique and the necessity for precise ventilatory parameter settings. Inappropriate settings can lead to suboptimal ventilation and oxygenation, increasing the risk of complications such as pneumothorax.

This review underscores the need for further research to optimize HFJV application and better understand its long-term clinical impacts. The insights gained provide valuable guidance for future clinical use.

**Key Message:** This systematic review examines the safety and efficacy of high frequency jet ventilation in adult and paediatric patients. The findings indicate that HFJV offers advantages in complex procedures, such as airway surgery and intensive care, by providing lower alveolar pressures and improved surgical conditions without significantly increased complication rates compared to conventional ventilation. However, HFJV requires precise parameter controls to avoid complications such as pneumothorax. Further research is needed to optimise its long-term clinical effects.

## 1 Introduction

Jet ventilation is a specific variant of mechanical ventilation that plays a central rolein airway and thoracic surgery, in imaging guided interventions as well as in emergency medicine [1]. However, this ventilation technique has not yet widely adopted in medical practice [2]. A possible reason for this could be the lack of specific recommendations from the relevant national scientific medical societies. Although jet ventilation is considered a suitable ventilation method, and clear guidelines exist for high frequency jet ventilation (HFJV) [3, 4], uncertainty and hesitation among healthcare professionals hinders the dissemination of this method [5]. Another reason may be the unfamiliarity with the underlying technique and the lack of manual access to the system, as compared to the use of a hand-bag breathing circuit in conventional ventilation.

Additionally, existing studies, which may partially be outdated, might not sufficiently demonstrate the clear advantages of HFJV, potentially affecting its adoption as a standard method for certain clinical procedures [2].

Jet ventilation, and in particularl high frequency jet ventilation (HFJV), represents a significant advancement in ventilation technology and is gaining increasing importance in modern medicine. Originally developed in the 1950s to improve ventilation during rigid bronchoscopy, this method was substantially refined by Douglas Sanders in 1967. Sanders’ introduction of a specialised adapter enabled continuous patient ventilation during surgical procedures via the bronchoscope by delivering intermittant high-pressure oxygen insufflation. By this measure, ambient air entrains the delivered oxygen portions, thereby increasing tidal volume without requiring a pressure driven closed ventilation system [1]. In the 1970s, HFJV further evolved, characterised by the use of high ventilation frequencies and small tidal volumes, typically smaller than the anatomical dead space [1].

High frequency ventilation (HFV) includes three main variants:

1. High frequency percussive ventilation (HFPV),
2. High frequency oscillatory ventilation (HFOV),
3. High frequency jet ventilation [6].

### High frequency percussive ventilation

This modality operates within a frequency ranging from 60 to 1.000 insufflations per minute [6].

HFPV combines rapid, pulsatile air jets via traditional ventilation cycles, allowing for enhanced gas distribution within the lungs [7]. It usually utilises smaller, more frequent volume pulses that are able to penetrate deeper lung regions, potentially reopening collapsed areas. The high frequency and low volumes are intended to reduce the risk of atelectasis, lung overdistension and other ventilation-associated lung injuries [7].

Another advantage of HFPV is its lower sedation and paralysis requirements, making it effective for bronchjo-pulmonary secretion clearance as well [8].

HFPV is frequently used in the treatment of severe respiratory disorders, such as Acute Respiratory Distress Syndrome (ARDS), and is also applied in cases of acute lung failure following smoke inhalation or burn injuries, in both paediatric and adult patients [7, 9].

### High frequency oscillatory ventilation

This ventilation technique operates at frequencies ranging from 180 to 1.200 cycles per minute [6].

HFOV combines low tidal volumes with a constant mean airway pressure and high breathing frequencies to optimise oxygenation and ventilation. This approach avoids the traumatic inflation and deflation of the lungs, which can occur with conventional ventilation [10].

HFOV utilises a simple circuit system to generate small tidal volumes while maintaining stable airway pressure. This helps to prevent lung damage caused by overdistension and volume overload, while achieving effective ventilation even with tidal volumes below the dead space [10].

HFOV is primarily used in patients with severe ARDS, especially when conventional ventilation methods prove to be insufficient [8] or when there is an increased risk of ventilator-induced lung injury [10].

### High frequency jet ventilation

High frequency jet ventilation operates at frequencies ranging from 100 to 1.500 breaths per minute [6]. This specialised ventilation technique delivers small gas volumes at high frequencies through narrow nozzles into the airway, achieving effective ventilation without generating high alveolar pressures [11].

Thanks to these mechanisms, HFJV is particularly suitable for patients requiring precise control of ventilation, such as in neonatal and paediatric intensive care, as well as during airway or thoracic surgeries [1]. It enables minimal movement in the surgical field, making it especially useful when conventional ventilation methods reach their limits or when gentle ventilation is required [12]. HFJV is also a helpful option in emergency situations where conventional ventilation methods are not applicable [1] such as in upper airway injury and inability to obtain an airtight sealing with a tracheal tube.

Modern jet ventilators use magnetic or fluidic valves to interrupt a continuous gas flow into precise pulses that transport the breathing gas into the lungs [1]. Built-in alarms and automatic shut-off functions provide protection against excessive airway volumes and pressures [1]. These devices are also equipped with humidification and warming features to prevent the patient’s airways from drying out and cooling down, and in particular to avoid mucosal damage such as “necrotizing tracheobronchitis” [1].

When delivering oxygen-air mixtures, modern HFJV allows for the adjustment of the fraction of inspired oxygen (FiO₂). However, the actual oxygen level reaching the patient is lower than its original setting at the ventilator. This phenomenon is called entrainment, and its impact is influenced by the amount of ambient air that flows inward alongside the oxygen. HFJV operates in the sense of a time-cycled, pressure-limited ventilation, meaning that changes in lung or chest wall compliance can impact minute ventilation [1].

Despite the challenges posed by low tidal volumes and high frequencies, effective gas exchange in HFJV is facilitated by specific mechanisms that are not present in conventional ventilation. These mechanisms are crucial to supporting gas exchange under these conditions [1].

A key mechanism of high frequency jet ventilation is **laminar flow**, particularly in small airways with a low Reynolds number. The velocity profile of this gas flow is parabolic: the flow velocity is highest in the centre of the airway and decreases towards the surrounding airway walls. This results in a rapid, axial gas flow in HFJV, while gas passively moves outward along the edges [1].

Another important mechanism is **Taylor dispersion**, which occurs through the interaction between axial parabolic velocity distribution and radial concentration gradients, enhancing molecular diffusion. This dispersion promotes additional mixing of gases, especially in the smaller airways, thereby facilitating effective gas exchange. In larger airways, radial gas-mixing is promoted by turbulent flow. These turbulences create forward-moving gas, contributing to mixing in a manner similar to Taylor dispersion.

Another significant mechanism in HFJV is **pendelluft** or collateral ventilation, resulting from regional differences in airway resistance and compliance. These differences cause some lung areas to fill or empty faster than others, producing a phase shift between neighbouring lung regions. This allows gas to move from one alveolus to another, promoting gas exchange but also increasing effective dead space ventilation. In addition to these flow mechanisms, **molecular diffusion** and **cardiogenic mixing** also contribute to gas exchange. Cardiogenic mixing results from the mechanical movement of the lungs caused by the heart’s nearby motion [1].

The key parameters in high frequency jet ventilation are frequency, working pressure, inspiratory time, and FiO₂. Tidal volume is not directly set but is a result of these parameters and the physical properties of the patient’s respiratory system. Adjusting these settings can influence CO₂ elimination and oxygenation, however, inappropriate settings may result in insufficient CO₂ exhalation or inadequate oxygenation [1].

HFJV can be applied via tracheal tubes, jet cannulas integrated in suspension laryngoscopes or specialised jet catheters, which can be positioned above or below the vocal cords. Thus, we speak of supraglottoc or infraglottic jet ventilation. These catheters often feature multiple lumens for functions such as intratracheal pressure monitoring or CO₂ sampling [1].

Settings for peak inspiratory pressure limits (PIP) and frequency are individually adjusted to ensure optimal ventilation and oxygenation while minimising the risk of barotrauma or other lung injuries [12, 1].

An important albeit grossly neglected feature in HFJV is gas conditioning. This comprises both: warming and humidifying the insufflated gas. Although there prevails a false belief that in relatively short procedures, gas conditioning might be not essential, the authors are convinced that for any duration of HFJV application, the physically maximal possible gas conditioning should always be applied. Warming the gas means elevating its temperature as close as possible to 37°C, but avoiding higher temperatures that may harm the airway mucosa. Concerning moistening, the maximally achievable relative humidity of the warmed gas should be aimed for. Due to physical limitations, some of the water vapor may condensate along the jet line, thus delivering water droplets instead of humidity. This might be relevant in very small patients such as neonates, who would be subjected to water intoxication. In adults this issue is negligeable. Modern jet ventilators automatically adjust the heating energy and water vaporizing to the delivered jet gas. Only in rare cases such as in microlaryngoscopic laser-light interventions, the gas humidity might cause a fogging of the microscope’s mirror. In this case, the conditioning must be reduced as far as necessary but maintained as high as possible.

The goal of this systematic review is to comprehensively review and synthesise safety measures and clinical outcomes of high frequency jet ventilation. Data from various relevant sources are analysed to illuminate specific aspects of HFJV. The review covers its use in both adult and paediatric medicine.

The analysis focuses on the methodological quality of the included studies to assess the reliability and applicability of the available evidence on the safety and effectiveness of HFJV. The extent to which different study designs, patient cohort selection, and reported clinical endpoints influence the interpretability of results and their relevance for clinical practice is examined.

This systematic approach aims to develop an evidence-based recommendation for the use of HFJV in specific medical contexts. Potential safety risks and the advantages and disadvantages of this technology for different medical disciplines are also discussed to provide a solid foundation for assessing the effectiveness and risks of HFJV.

## 2 Methods

### 2.1 Literature research outcome

The database search identified 4,403 potentially relevant publications in PubMed, Cochrane Library, and Livivo. After removing 4,072 duplicates, the abstracts of the remaining 331 articles were screened for potential suitability in a systematic review. In this process, 183 articles were excluded because they did not meet the basic inclusion criteria or appeared to be of unclear relevance.

The remaining 148 articles were further screened based on their titles for relevance to the review’s objective, resulting in further exclusion of 85 articles. This was followed by an abstract screening, which led to the exclusion of nine additional articles. The remaining 57 articles underwent a full-text analysis. During this step, six articles were excluded due to limited direct relevance to the study’s objective. Another eight studies were excluded because they did not meet the established criteria for high frequency jet ventilation, since in these studies, the set respiratory rate was below 100 breaths per minute, thus not meeting the requirements for HFJV [6].

The PRISMA Search Reporting Extension Checklist (PRISMA) was applied to conduct this systematic review. The literature review process is presented in Figure 1, which displays the PRISMA flow diagram and divides the process into the three main phases of study identification, screening, and inclusion [13].

**Figure 1:**
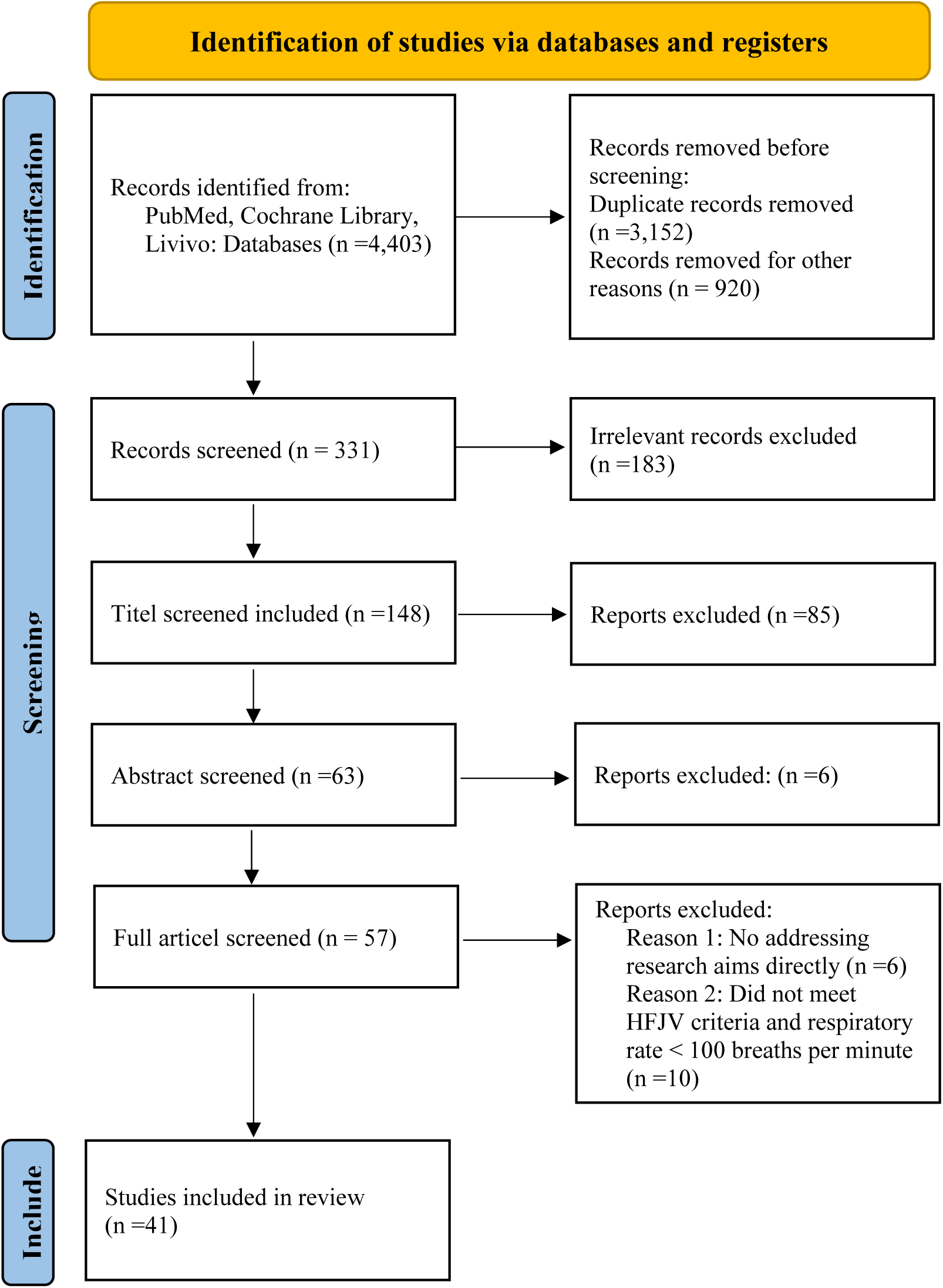
PRISMA flow diagram From: Page MJ, McKenzie JE, Bossuyt PM, Boutron I, Hoffmann TC, Mulrow CD, et al. The PRISMA 2020 statement: an updated guideline for reporting systematic reviews. BMJ 2021;372:n71. doi: 10.1136/bmj.n71 [13].

### 2.2 Literature research methods

For the systematic review, a scientifically grounded methodology was applied in accordance with the established PRISMA guidelines (Preferred Reporting Items for Systematic Reviews and Meta-Analyses). This model provides proven guidelines for the structured planning, execution, and documentation of systematic reviews, ensuring a transparent and traceable approach [13].

**Table 1:** Search Terms for Literature Review.

| Search terms | PubMed | Cochrane Library | LIVIVO |
| --- | --- | --- | --- |
| Jet Ventilation | Jet Ventilation | Jet Ventilation | Jet Ventilation |
| HFJV | HFJV | HFJV | HFJV |
| HFJV and Physical | HFJV and Physical | HFJV and Physical | HFJV and Physical |
|  | HFJV and Physical | HFJV and Physical | HFJV and Physical |
| HFJV not HFOV | HFJV, not HFOV | HFJV, not HFOV | HFJV, not HFOV |
| High-Frequency Jet Ventilation | High-Frequency Jet Ventilation | High-Frequency Jet Ventilation | High-Frequency Jet Ventilation |
| “High Frequency Jet Ventilation“ | “High Frequency Jet Ventilation“ | “High Frequency Jet Ventilation“ | “High Frequency Jet Ventilation“ |

The systematic literature review was based on a precise search strategy to comprehensively capture relevant articles. Boolean operators such as “and” and “not” were utilised (Table 1) to effectively refine the search results and exclude irrelevant articles. The “and” operator was used to narrow down results to studies containing multiple key terms, while “not” served to exclude unrelated fields, thereby maximising the precision of the results [14]. To ensure the relevance and currency of the studies, clear inclusion and exclusion criteria were also defined (Table 2). A further crucial aspect was the careful selection of search terms, as insufficiently precise ones could overlook potentially relevant studies. Including synonyms and -specific keywords optimised the relevance of the search strategy [15].

To strike a balanced approach between specificity and sensitivity in the search results, test runs were conducted, and the search terms were continuously refined.

**Table 2:** Inclusion and exclusion criteria of the literature search.

| Inclusion criteria | Exclusion criteria |
| --- | --- |
| High frequency jet ventilation | Studies older than 2001 |
| Application in Pediatrics and Adult Medicine | Terms: HFV, HFOV, HFPV |
| Studies written in English and German | General journal articles, comments, opinion pieces, and newsletters, retraction of publications, withdrawn publications |
|  | Respiratory rate of patients below 100 breaths per minute |

### 2.2 Quality appraisal

The quality assessment of the clinical studies is based on the medical level of evidence [16].

**Table 3:** Quality assessment of clinical studies based on the medical level of evidence.

| Definition/Abbreviation | Description |
| --- | --- |
| **Quality** | The quality assessment of the clinical studies is based on the level of evidence from the Oxford Centre for Evidence-Based Medicine (OCEBM) |
| **I** | **Systematic review of randomized trials**<br>Systematic reviews or meta-analyses of randomized controlled trials (RCTs) |
| **II** | **Randomized trial**<br>Individual randomized controlled trial or systematic reviews of cohort studies |
| **III** | **Non-randomized controlled cohort/ follow-up study**<br>Single cohort study, low-quality RCTs or systematic reviews of case-control studies, prospective studies with pre-specified inclusion criteria |
| **IV** | **Case-series, case-control, or historically controlled studies**<br>Case series, case reports, or single case-control study |
| **V** | **Mechanism-based reasoning**<br>Expert opinion without explicit critical appraisal |

## 3 Results

### 3.1 Summary of results

In this section, the key findings of the analysed studies are summarised, with each study evaluated based on a Quality Score. The references are presented in a structured manner and serve as the basis for the subsequent discussion.

#### 3.1.1 High Frequency Transcricoid Jet Ventilation Prior to Neck Surgery—A 10-Year Retrospective Case Series Analysis

Kisch-Wedel HL, Reichert B, Niedermeyer HP.

Year: 2020.

Study-design: Retrospective analysis.

Impact Factor: 0.38 (2020).

Patient collective: Total number of patients: 39,477 who underwent neck surgery. Patients managed with TCHFJV: 1,489 (3.8% of the total patients). Gender: 1,090 males, 399 females. Age: 62 ± 13 years. BMI: 25 ± 0.2 kg/m². ASA class: 3 ± 0.5. Study period: 10 years.

Methods: TCHFJV (Transcutaneous high frequency jet ventilation) was applied before anesthesia induction in awake, spontaneously breathing patients. These patients had a suspected or known difficult airway and were scheduled for head and neck surgery.

**Outcome: TCHFJV was successful in 1,479 (99%) patients, with endotracheal intubation performed in 93% of cases. Complications occurred in 1.5% of cases, the majority being non-severe.**

Quality Score: 4

#### 3.1.2 Effects of helium on high frequency jet ventilation in a rabbit model of respiratory failure

Buczkowski PW, Fombon FN, Russell WC, Young JD, Jordan S, Howells PA, Grounds RM.

Year: 2005.

Study-design: Experimental study.

Impact Factor: 6.880 (2023).

Patient collective: Trachea-lung model.

Methods: HFJV was applied at varying degrees of stenosis (2.5–8.5 mm) via supraglottic, transglottic, and infraglottic routes. Pressures and gas volumes were measured. Measurements of distal tracheal pressures were repeated for each route at steady state for each stenosis diameter using both 100% oxygen and helium–oxygen (50% oxygen, 50% helium). The ventilator’s output was measured during operation on oxygen and helium–oxygen.

**Outcome: Using a helium-oxygen mixture during HFJV resulted in an 18% increase in minute ventilation volume at the same or lower airway pressures compared to 100% oxygen. Significant pressure reductions were observed at stenosis diameters less than 4 mm.**

Quality Score: 4

#### 3.1.3 Use of high-frequency jet ventilation for percutaneous transhepatic cholangioscopy in patients with complicated biliary diseases

Denys A, Lachenal Y, Duran R, Moutardier V, Giovannini M.

Year: 2014.

Study-design: Prospective single-center study.

Impact Factor: 2.082 (2014).

Patient collective: 51 patients, 41 treated with HFJV and 10 with conventional ventilation. Gender: 14 women. Mean age: 66 years. Tumors: 66 tumors in total, including 56 hepatic tumors, 5 pulmonary tumors, and 5 renal tumors. Median tumor size: 16 ± 8.7 mm.

Methods: The study examined the feasibility and potential benefits of HFJV in tumor ablation techniques for the liver, kidneys, and lungs. Data on anesthesia and procedure duration, as well as mean CO₂ levels, were recorded. CT and ultrasound guidance were used for tumor ablation procedures. Organ movements during HFJV were measured.

**Outcome: HFJV was feasible in 80% of patients, allowing near immobility of internal organs during tumor ablation, with an average organ movement of 0.3 mm in the x and y axes and less than 3.75 mm in the z axis. No specific complications or hypercapnia related to HFJV were observed.**

Quality Score: 2

#### 3.1.4 Einseitige High-Frequency-Jet-Ventilation als Ergänzung der Ein-Lungenbeatmung bei thoraxchirurgischen Eingriffen

Knüttgen D, Zeidler D, Vorweg M, Doehn M.

Year: 2001.

Study-design: Case study.

Impact Factor: 0.672 (2001).

Patient collective: Two patients with significant functional limitations of the left lung.

Methods: The left lung was ventilated using IPPV, with tidal volume reduced to 200 ml in the first patient due to high airway pressures. The right lung was simultaneously ventilated using HFJV.

**Outcome: Both patients exhibited stable arterial oxygen saturation (96-100%) and normal CO2 partial pressure (45 mmHg) during and after the operation, with no complications reported. This innovative ventilation technique facilitated effective oxygenation and CO2 elimination despite the patients’ impaired lung function.**

Quality Score: 4

#### 3.1.5 Mortality Risk Factors in Preterm Infants Treated with High-Frequency Jet Ventilation

Wheeler CR, Stephens H, O’Donnell RM.

Year: 2020.

Study-design: Retrospective cohort study.

Impact Factor: 2.066 (2020).

Patient collective: 53 preterm infants (<34 weeks gestational age, <2000 g birth weight), excluded if they received HFJV at admission, included if they failed conventional ventilation.

Methods: Demographic data of the subjects as well as ventilation parameters and laboratory results were extracted and analyzed. Univariate and multivariate logistic regression analyses were used to evaluate variables predicting mortality. Receiver Operating Characteristic (ROC) curves were constructed to assess the predictive accuracy of the multivariate risk score.

**Outcome: Identified independent predictors of mortality: female sex, closed ductus arteriosus, and oxygen saturation index > 5.5 after 4 hours of HFJV. These factors were used to create a multivariate risk score, which demonstrated high predictive accuracy for mortality. The study emphasizes that OSI is a useful non-invasive measure for guiding interventions and determining risk stratification in premature infants.**

Quality Score: 3

#### 3.1.6 Transcutaneous monitoring of partial pressure of carbon dioxide in adults using high-frequency jet ventilation

Weiss E, Dreyfus JF, Fischler M, Le Guen M.

Year: 2017.

Study-design: Prospective, observational, single-centre study.

Impact Factor: 4.164.

Patient collective: 83 patients: 56 male (67.5%) and 27 female (32.5%), median age of 57 years (range 47.5–66.5 years), median weight of 67.5 kg (range 39–140 kg), median height of 168 cm (range 151–185 cm), ASA1 (1.2%), ASA2 (50%), ASA3 (42.5%), and ASA4 (6.2%). Underlying conditions included cancer (42.2%), lung transplantation (26.5%), benign lesions (16.9%), and other conditions (14.5%).

Methods: Anaesthesia with propofol, remifentanil, and succinylcholine or atracurium. PtCO2 measurements using a heated transcutaneous ear sensor every 10 min, compared with PaCO2 values.

**Outcome: The aim of the study was to determine the accuracy of transcutaneous CO2 measurement (PtCO2) in patients undergoing therapeutic rigid bronchoscopy under HFJV. PtCO2 monitoring was found to be highly correlated with PaCO2 (bias of 4.0 ± 0.56 mmHg). PtCO2 effectively detected dangerous hypercapnic situations, though larger limits of agreement were observed at higher PaCO2 levels.**

Quality Score: 3

#### 3.1.7 Transtracheal high frequency jet ventilation for endoscopic airway surgery: a multicentre study

Bourgain JL, Desruennes E, Fischler M, Ravussin P.

Year: 2001.

Study-design: Prospective multicenter study.

Impact Factor: 2.145.

Patient collective: 643 patients (523 males, 120 females, 632 adults, 11 children), ages 1 month to 86 years, underwent laryngoscopy or laryngeal laser surgery, received transtracheal HFJV. Exclusion criteria: infection at the puncture site, coagulation disorders. Duration: 1 year conducted in three different hospitals.

Methods: Catheter inserted through cricothyroid membrane, HFJV started with specific settings, procedures under general anesthesia, complications and outcomes recorded.

**Outcome: HFJV allowed excellent surgical visibility and adequate ventilation. Complications were rare but included subcutaneous emphysema (8.4%) and pneumothorax (1%). Arterial oxygen desaturation was more frequent during laser procedures and in overweight patients. No catheter was misplaced or caused bleeding.**

Quality Score: 3

#### 3.1.8 The Veres Adapter: Clinical Experience with a New Device for Jet Ventilation via a Laryngeal Mask Airway During Flexible Bronchoscopy

Veres J, Slavei K, Errhalt P, Seyr M, Ihra G.

Year: 2011.

Study-design: Clinical study.

Impact Factor: 5.7.

Patient collective: 30 adults (age ≥18 years, weight ≤120 kg), average age: 59 years, average weight: 71 kg (without COPD) and 86 kg (with COPD), average height: 166 cm (without COPD) and 169 cm (with COPD).

Methods: The Veres adapter was connected to a size 4 or 5 laryngeal mask airway (LMA) and HFJV was performed. Oxygen saturation, transcutaneous carbon dioxide, supraglottic airway pressure, and hemodynamic data were recorded and analyzed. Bronchoscopy was performed using flexible bronchoscopes, and a lidocaine solution was used for topical anesthesia.

**Outcome: The Veres adapter enables effective HFJV during bronchoscopy, resulting in rapid recovery and stable hemodynamic conditions. The adapter could be a beneficial alternative to the endotracheal tube, as it better maintains the airway with a larger diameter. Short procedure times and fast recovery were observed. Mild hypercapnia was the most common minor adverse effect.**

Quality Score: 3

#### 3.1.9 The quality of the surgical field during functional endoscopic sinus surgery—The effect of the mode of ventilation—A randomized, prospective, double-blind study

Chhetri DK, Berke GS, Gerratt BR.

Year: 2009.

Study-design: Prospective, randomized, double-blind study.

Impact Factor: 2.78.

Patient collective: 22 patients (6 women, 16 men), age 24 to 68 years, ASA I-II patients with chronic rhinosinusitis, with or without nasal polyps. Exclusion criteria: patient refusal, diabetes, coagulation disorders, chronic restrictive and obstructive pulmonary disease, treatment with antiplatelet aggregation and anticoagulation medication.

Methods: Patients were randomly assigned to either the HFJV or IPPV group. Both groups received the same general anesthesia. HFJV: 150/min, I ratio 1:1, gas pressure 2–2.5 bar, IPPV: 10 ml/kg gas, frequency 10/min, I ratio 1:2. The quality of the surgical field was assessed and the total blood loss was measured. Two groups: Group 1 ventilated by IPPV, Group 2 ventilated by HFJV.

**Outcome: The study investigated the effects of HFJV compared to intermittent positive pressure ventilation (IPPV) in patients undergoing functional endoscopic sinus surgery (FESS). The mean airway pressure was significantly lower in the HFJV group. Blood loss in the HFJV group was 170 cc, and in the IPPV group 318.18 cc. The quality of the surgical field was better in the HFJV group.**

Quality Score: 2

#### 3.1.10 The effects of high-frequency jet ventilation (HFJV) on pneumoperitoneum-induced cardiovascular changes during laparoscopic surgery

Bickel A, Trossman A, Kukuev I, Eitan A.

Year: 2011.

Study-design: Randomized prospective trial.

Impact Factor: 3.1 (2023).

Patient collective: 25 healthy patients scheduled for elective laparoscopic cholecystectomy, ASA I–II. Control group: 12 patients under conventional mechanical ventilation. Study group: 13 patients under HFJV. Randomization: Sealed envelope method.

Methods: Cardiac functionality was continuously evaluated using arterial pressure wave analysis with Edwards Flo-Trac sensor and Vigileo monitor.

**Outcome: The study investigated the influence of HFJV on cardiovascular changes during laparoscopic cholecystectomy. HFJV showed a reduction in the negative cardiovascular effects compared to conventional mechanical ventilation.**

Quality Score: 2

#### 3.1.11 Rescue high-frequency jet ventilation versus conventional ventilation for severe pulmonary dysfunction in preterm infants

Rojas-Reyes MX, Orrego-Rojas PA.

Year: 2015.

Study-design: Multi-centre trial, enrolment between January 1987 and March 1989.

Impact Factor: 12.008 (2023).

Patient collective: 166 preterm infants younger than 7 days of age and weighing ≤ 750 grams at birth, with pulmonary interstitial emphysema. Randomization was performed centrally through a 24-hour hotline call. Exclusions: 22 out of 166 infants were excluded after initial randomization.

Methods: HFJV with 400 to 450 cycles/min (treatment group), conventional ventilation (CV) with rates of 60 to 100 breaths/min, short inspiratory time (control group). 70 infants were assigned to CV, and 74 infants to HFJV. Cross-over to alternate therapy was allowed if infants failed allocated ventilator therapy.

**Outcome: The study compared the use of HFJV with conventional ventilation in premature infants with severe lung dysfunction. No significant differences in overall mortality. HFJV showed lower mortality before cross-over. No significant differences in CLD, new air leaks, IVH (grades 3 and 4), necrotizing tracheobronchitis, or airway obstruction.**

Quality Score: 2

#### 3.1.12 Rapid pacing and high-frequency jet ventilation additively improve catheter stability during atrial fibrillation ablation

Aizer A, Qiu JK, Cheng AV, Wu PB, Barbhaiya CR, Jankelson L, Linton P, Bernstein SA, Park DS, Holmes DS, Chinitz LA.

Year: 2020.

Study-design: Randomized controlled trial with crossover design.

Impact Factor: 3.1 (2023).

Patient collective: 40 patients undergoing ablation for paroxysmal atrial fibrillation.

Methods: 40 patients were enrolled and received ablation lesions at 15 prespecified locations. They were randomly assigned to undergo rapid atrial pacing for either the first or second half of each lesion. Within each group, half of the patients received HFJV and the other half standard ventilation. Standard deviation of contact force was the primary endpoint to examine contact force variability.

**Outcome: The study investigates the effects of rapid cardiac pacing and HFJV on catheter stability during ablation of atrial fibrillation. Lesions with both pacing and HFJV had the greatest reduction in contact force standard deviation (4.35 ± 2.81 g), confirming an additive benefit. Simultaneous pacing and HFJV further improves catheter stability over pacing or HFJV alone to optimize ablation lesions.**

Quality Score: 2

#### 3.1.13 Jet Ventilation Reduces Coronary Sinus Movement in Patients Undergoing Atrial Fibrillation Ablation: An Observational Crossover Study

**Jet Ventilation Reduces Coronary Sinus Movement in Patients Undergoing Atrial Fibrillation Ablation: An Observational Crossover Study**

Maeyens C, Nokerman P, Casado-Arroyo R, Abugattas De Torres JP, Alexander B, Engelman E, Schmartz D, Tuna T.

Year: 2023.

Study-design: Prospective, interventional, single-center, crossover study.

Impact Factor: 4.945 (2023).

Patient collective: 21 patients (18 men, 3 women), median age: 59 years (Q1-Q4: 55–70), median weight: 84 kg (Q1-Q4: 71–92), median height: 1.78 m (Q1-Q4: 1.72–1.82), median BMI: 26.3 kg/m² (Q1-Q4: 24.0–28.0). Inclusion criteria: patients ≥ 18 years, scheduled for atrial fibrillation ablation, BMI ≤ 30 kg/m², ASA score 1–3. Exclusion criteria: refusal to participate, pre-existing lung disease, heart failure (LVEF < 30%). Sample size: 15 patients required for significance, 21 patients included to account for a 25% dropout rate.

Methods: Each patient was ventilated with both normal mechanical ventilation and HFJV. During each ventilation mode, displacements of the cardiac structure were measured by the EnSite Precision mapping system using a catheter placed in the coronary sinus.

**Outcome: The study investigates the effects of HFJV compared to conventional mechanical ventilation (IPPV) in patients undergoing atrial fibrillation ablation. The results showed a significant reduction in coronary sinus movement when using HFJV compared to conventional ventilation. Median displacement: 2.0 mm (HFJV) vs. 10.5 mm (IPPV).**

Quality Score: 3

#### 3.1.14 Is HFJV a better alternative ventilation technique for percutaneous dilatational tracheostomy? A randomized trial

Altinsoy S, Sayin MM, Özkan D, Çatalca S, Ergil J.

Year: 2022.

Study-design: Randomized controlled trial.

Impact Factor: 2.508 (2023).

Patient collective: 75 patients, Group ETT (Endotracheal Tube): N=25, Group LMA (Laryngeal Mask Airway): N=25, Group HFJV: N=25.

Methods: Patients were randomized into one of the three groups with computer-generated random numbers. Demographic data, duration of PDT, complications such as ETT cuff puncture and tube transaction, accidental extubation, difficult cannula insertion, bleeding, desaturation during the procedure, and arterial blood gases immediately before and after the procedure were recorded.

**Outcome: The study investigates the use of HFJV compared to conventional ventilation methods (LMA and ETT) during PDT. The PDT duration was significantly shorter in the HFJV group compared to the other groups. Fewer complications were observed in the HFJV group.**

Quality Score: 2

#### 3.1.15 Initiation of a High-Frequency Jet Ventilation Strategy for Catheter Ablation for Atrial Fibrillation

Sivasambu B, Hakim JB, Barodka V, Chrispin J, Berger RD, Ashikaga H, Ciuffo L, Tao S, Calkins H, Marine JE, Trayanova N, Spragg DD.

Year: 2018.

Study-design: Prospective cohort study.

Impact Factor: 5.524 (2023).

Patient collective: 285 patients, including 84 HFJV and 84 control patients. Age: HFJV 63.1 years, control 61.1 years (p = 0.237). Gender: Female HFJV 34.5%, control 26.2% (p = 0.240). BMI: HFJV 30.3 kg/m², control 31.1 kg/m² (p = 0.303). Period: 2015 to 2017.

Comorbidities: CHF: HFJV 7.1%, control 16.7% (p = 0.057), DM: Both groups 17.9% (p = 1.000), CVA: HFJV 11.9%, control 7.1% (p = 0.293), HTN: HFJV 57.1%, control 63.1% (p = 0.431), OSA: HFJV 16.7%, control 23.8% (p = 0.249).

Methods: Patients undergoing pulmonary vein isolation (PVI) at Johns Hopkins Hospital were prospectively enrolled in a registry. Patients were supported either by standard ventilation or HFJV. Key parameters included procedural characteristics, adverse event rates, and 1-year outcomes.

Outcome: The study investigated the use of HFJV during PVI in atrial fibrillation (AF). Atrial arrhythmia recurrence at 1 year was 31% in HFJV patients vs. 50% in control patients. HFJV was associated with improved catheter stability and contact force adequacy.

Quality Score: 3

#### 3.1.16 High-Frequency Jet Ventilation Improves Gas Exchange in Extremely Immature Infants with Evolving Chronic Lung Disease

Sivasambu B, Hakim JB, Barodka V, Chrispin J, Berger RD, Ashikaga H, Ciuffo L, Tao S, Calkins H, Marine JE, Trayanova N, Spragg DD.

Year: 2006.

Study-design: Prospective observational.

Impact Factor: 5.524 (2023).

Patient collective: 10 mechanically ventilated extremely immature infants with evolving CLD; median gestational age 23.6 weeks (range 22+3/7 to 26+3/7), median birth weight 650 g (range 390 to 1020 g). Chorioamnionitis: Histologically confirmed in all patients. Period: 2002 to October 2004.

Methods: Chorioamnionitis was confirmed in all infants, patent ductus arteriosus was ligated in five patients, and Ureaplasma urealyticum was cultured from the trachea in four patients. HFJV was initiated when oxygenation index > 10 or exhaled tidal volume ≤ 7 mL/kg were required to maintain PaCO2 < 60 mmHg. Ventilatory stabilization and weaning from mechanical ventilation with extubation to nasal CPAP were the goals. Adjustments were made to pressure amplitude to keep PaCO2 between 45 and 55 mmHg.

**Outcome: PaCO2 decreased within 1 hour of HFJV initiation and oxygenation index decreased by 24 hours. 9 out of 10 patients survived, and all were successfully extubated to nasal CPAP in a median of 15.5 days. Gas exchange and ease of extubation improved.**

Quality Score: 3

#### 3.1.17 Comparison of superimposed high-frequency jet ventilation with conventional jet ventilation for laryngeal surgery

Leiter R, Aliverti A, Priori R, Staun P, Lo Mauro A, Larsson A, Frykholm P.

Year: 2012.

Study-design: Prospective, randomized clinical comparative study.

Impact Factor: 6.880 (2023).

Patient collective: 16 patients undergoing minor laryngeal surgery. Gender: 7 males, 9 females. Age range: 33 to 72 years. Inclusion criteria: Age >18 years, ASA classification I–II, BMI <35 kg/m². Exclusion criteria: Respiratory diseases (e.g., asthma, COPD), symptomatic airway stenosis.

Methods: Four different JV techniques were applied in random order for 10-min periods: SHFJVSG (superimposed high-frequency jet ventilation, supraglottic), NFJVSG (normal frequency jet ventilation, supraglottic), HFJVSG (high-frequency jet ventilation, supraglottic), and HFJVIG (high-frequency jet ventilation, infraglottic). Chest wall volume variations were continuously measured with opto-electronic plethysmography (OEP), intratracheal pressure was recorded, and blood gases were measured.

**Outcome: SHFJVSG was associated with increased end-expiratory chest wall volume and tidal volume compared with the other JV modes. All modes provided adequate oxygenation and ventilation.**

Quality Score: 2

#### 3.1.18 High frequency jet ventilation through mask contributes to oxygen therapy among patients undergoing bronchoscopic intervention under deep sedation

Yang M, Wang B, Hou Q, Zhou Y, Li N, Wang H, Li L, Cheng Q.

Year: 2021.

Study-design: Prospective randomized cohort study.

Impact Factor: 2.688 (2021).

Patient collective: 161 patients (150 after exclusions). COT Group: 50 patients (FiO2 of 1.0, 12 L/min). NFJV Group: 50 patients (FiO2 of 1.0, driving pressure of 0.1 MPa, respiratory rate (RR) of 15 bpm). HFJV Group: 50 patients (FiO2 of 1.0, driving pressure of 0.1 MPa, RR of 1200 bpm). Registration: Chinese Clinical Trial Registry (ChiCTR2000031110). Inclusion criteria: Scheduled for electric flexible bronchoscopy, duration of operation: 20 to 60 minutes, age: 18–80 years. Exclusion criteria: Diagnosed with cardiac respiratory failure and coma, T-tube, endotracheal intubation, tracheotomy, or SpO2 < 90% in room air before surgery, history of mental and neurological disorders, sedative or hypnotic drugs, and alcohol abuse.

Methods: Anesthesia procedure: Monitoring with electrocardiogram (ECG), pulse oximetry (SpO2), and blood pressure. Local anesthesia with lidocaine (1%, 10 ml). Use of an endoscopic face mask for oxygenation. Parameters recorded included pulse oxygen saturation (SpO2), mean arterial blood pressure, heart rate, and arterial blood gas. Analysis of arterial blood gases 15 minutes after the procedure began. Documentation of procedure duration, dose of anesthetics, and adverse events.

**Outcome: The study examines the effectiveness and safety of HFJV compared to conventional oxygen therapy (COT) and normal-frequency jet ventilation (NFJV) during bronchoscopic interventions under deep sedation. The results show that HFJV significantly improves patients’ oxygen levels (PaO2) without increasing the risk of hypercapnia.**

Quality Score: 2

#### 3.1.19 High frequency jet ventilation in preterm infants: experience from Western Australia

Venkataraman R, Kamlin O, Davis P, Roehr CC.

Year: 2018.

Study-design: Case-control study.

Impact Factor: 1.737 (2018).

Patient collective: 100 preterm infants admitted between February 2010 and June 2014, needing HFJV. HFJV Group (Cases): 50 preterm infants, Control Group: 50 preterm infants. Included infants born <32 weeks gestation, very low birth weight (VLBW <1500 g), and those at high risk for severe lung disease progressing to bronchopulmonary dysplasia (BPD). Exclusion criteria were congenital anomalies or critical cardiac conditions.

Methods: Controls were matched to cases (1:1) on gestation, birthweight, gender, place of birth, growth status, antenatal glucocorticoids, and dexamethasone as treatment for severe bronchopulmonary dysplasia (BPD). Logistic regression analysis was used to control for confounders. Data on all preterm infants who needed HFJV (Cases: n=50) and 50 controls during the study period were analyzed.

**Outcome: The study investigated whether HFJV in preterm infants reduces mortality or the need for home oxygen. HFJV was not significantly associated with reduced odds of death or discharge home on oxygen after adjusting for mean airway pressure and adjuvant therapy (Adjusted OR: 1.46, p=0.687). However, the odds of death before discharge were higher in the HFJV group (OR: 6.00, p=0.013).**

Quality Score: 4

#### 3.1.20 High Frequency Jet Ventilation during Initial Management, Stabilization, and Transport of Newborn Infants with Congenital Diaphragmatic Hernia: A Case Series

Choudhary S, Malhotra A, Lahra A, Barfield CA, Hunt RW.

Year: 2013.

Study-design: Retrospective chart review.

Impact Factor: 1.531 (2023).

Patient collective: 25 infants with antenatally diagnosed congenital diaphragmatic hernia (CDH). Males: 16 (64%), females: 9 (36%). Birth weight: Average 2868 grams. Gestational age: Average 37.2 weeks. Type of CDH: Left-sided 19 (78%), bilateral 1 (4%). Other congenital anomalies: 10 infants (40%), including congenital heart disease, micrognathia, small kidneys, and bilateral pelviectasis. Seven were preterm.

Methods: Retrospective chart review was performed for infants with antenatal diagnosis of CDH born between 2004 and 2009 at Mount Sinai Hospital, Toronto, Ontario, Canada. HFJV was initiated either as the initial mode or as rescue therapy. Blood gases were measured before and after HFJV initiation. Transport involved a specialized team and equipment to ensure stability. Equipment included Airborne brand portable transport incubators modified for HFJV, Life Pulse HFJV (Bunnell Inc.), and iNOvent for NO delivery.

**Outcome: The aim of the study was to assess whether HFJV reduces the likelihood of death or discharge with oxygen therapy in preterm infants. HFJV showed significant improvement in blood gases (PaCO2 and pH) and was safe and effective for transporting infants with CDH. Seven out of 25 infants survived to discharge.**

Quality Score: 4

#### 3.1.21 Electromagnetic navigation system combined with High-Frequency-Jet-Ventilation for CT-guided hepatic ablation of small US-Undetectable and difficult to access lesions

Ren F, Li Q, Gao X, Zhu K, Zhang J, Chen X, Yan X, Chu D, Hu L, Gao Z, Wu Z, Wu R, Lv Y.

Year: 2019.

Study-design: Prospective study.

Impact Factor: 3.589 (2019).

Patient collective: 21 patients (15 men, 6 women), mean age 67, with 27 lesions (14 hepatocellular carcinomas and 13 metastases), mean tumor size 12 ± 5.7 mm, all lesions invisible on US and difficult to reach on CT, requiring a double-oblique approach.

Methods: Electromagnetic navigation-based probe placement combined with HFJV was used for CT-guided hepatic ablation. Procedures involved placing the applicator near the tumor using EMN and performing ablation with HFJV to ensure stability and precision.

**Outcome: The study evaluated the safety and efficacy of using an electromagnetic navigation system combined with high-frequency jet ventilation for CT-guided ablation of difficult-to-access, ultrasound-invisible liver tumors. Complete ablation was achieved in 100% of cases at 3- and 6-month MRI follow-up. The applicator was accurately placed on the first pass in 96% of cases. There were no major complications reported.**

Quality Score: 3

#### 3.1.22 Die Hochfrequenz-Jet-Ventilation in der HNO-Heilkunde – chirurgische und anästhesiologische Aspekte

Helmstaedter V, Tellkamp R, Schwab B, Lenarz T, Durisin M.

Year: 2014.

Study-design: Retrospective chart review.

Impact Factor: 2.013 (2014).

Patient collective: 43 patients: 39 adults (15 men, 24 women, average age 59 years, range 19–88 years), 4 children (3 girls, 1 boy, ages 1, 3, 11). Number of procedures performed: 46 microlaryngoscopies and tracheoscopies. Study period: June 2012 to February 2013. Average BMI of adults: 27 kg/m² (range 17–42 kg/m²). ASA classification: ASA I: 24% (10 patients), ASA II: 54% (23 patients), ASA III: 22% (9 patients).

Methods: The jet catheters were usually inserted orally by the anesthesiologist or ENT surgeon using conventional Macintosh laryngoscopes and occasionally a Magill forceps. In some cases, nasal insertion through a Wendl tube was chosen to stabilize the catheter. During surgeries involving the use of CO₂ lasers, laser-resistant catheters and equipment were utilized to prevent burns and ensure patient safety. Continuous monitoring of ventilation was achieved through observation of thoracic excursions and auscultation.

**Outcome: The aim was to evaluate experience with HFJV for benign laryngotracheal lesions and to highlight relevant points for a safe and cooperative procedure. Two cases had reversible desaturations to 70%. In one case, emergency re-intubation was necessary due to desaturation below 50%. No other significant complications occurred.**

Quality Score: 4

#### 3.1.23 A multi-centre prospective random control study of superimposed high-frequency jet ventilation and conventional jet ventilation for interventional bronchoscopy

Wang T, Pei Y, Qiu X, Wang J, Wang Y, Zhang J.

Year: 2022.

Study-design: A multi-centre prospective random single-blind clinical trial.

Impact Factor: 1.152 (2022).

Patient collective: 60 patients. SHFJV Group (Trial Group): 29 patients, CHFJV Group (Control Group): 31 patients. Age: 18 to 70 years, requiring ventilation support during interventional bronchoscopy, voluntary participation with signed informed consent. Exclusion criteria: inability to withstand hyperextension or rotation of the neck, inability to undergo anesthesia in the supine position, acute exacerbation of asthma and COPD, or risk of pneumothorax, respiratory communicable diseases like tuberculosis and influenza, severe cardiovascular and cerebrovascular diseases, end-stage diseases, participation in other studies within the last 3 months, any contraindication to anesthesia.

Methods: Patients who underwent diagnostic or therapeutic bronchoscopy under general anesthesia were admitted and divided into two groups: SHFJV group (trial group) and CHFJV group (control group). PaO2 and PaCO2 were recorded before anesthesia, during, and after the procedure. SpO2 and etCO2 were recorded every 10 minutes throughout the procedure.

Patients were observed until 24 hours post-bronchoscopy.

**Outcome: The aim of the study was to investigate the efficacy and safety of superimposed high-frequency jet ventilation (SHFJV) compared to conventional high-frequency jet ventilation (CHFJV) during interventional bronchoscopy. SHFJV provided more effective and stable ventilation, especially in procedures longer than 90 minutes. The trial group showed more stable etCO2 levels and fewer instances of PaCO2 ≥ 50 mmHg compared to the control group. The PaO2 levels were higher in the control group during the procedure.**

Quality Score: 2

#### 3.1.24 The effects of high-frequency jet ventilation (HFJV) on pneumoperitoneum-induced cardiovascular changes during laparoscopic surgery

Bickel A, Trossman A, Kukuev I, Eitan A.

Year: 2011.

Study-design: Randomized prospective trial.

Impact Factor: 3.747 (2011).

Patient collective: 25 healthy patients scheduled for elective laparoscopic cholecystectomy (13 in the HFJV group, 12 in the CMV group). Age: Average 40.6 years in the CMV group and 39.5 years in the HFJV group. Gender: 3 men and 9 women in the CMV group, 6 men and 7 women in the HFJV group. Body Mass Index (BMI): Average 27.4 kg/m² in the CMV group and 27.3 kg/m² in the HFJV group. Medical history: No significant differences regarding ischemic heart disease (IHD), hypertension, diabetes, or pulmonary diseases.

Methods: Both groups were categorized as ASA I–II and underwent total intravenous anesthesia. Cardiac functionality was continuously evaluated by analysis of arterial pressure wave changes (Edwards Flo-Trac sensor and Vigileo monitor).

**Outcome: The aim of this study was to evaluate the benefits of HFJV to reduce the adverse cardiovascular effects during laparoscopic cholecystectomy. Significant reduction in cardiac output in the control group during the initiation of anti-Trendelenburg position, no significant hemodynamic changes under HFJV during the same condition. Total peripheral resistance increased during pneumoperitoneum in both groups.**

Quality Score: 2

#### 3.1.25 High-frequency jet ventilation for endolaryngotracheal surgery – chart review and procedure analysis from the surgeon’s and the anaesthesiologist’s point of view

Helmstaedter V, Tellkamp R, Majdani O, Warnecke A, Lenarz T, Durisin M.

Year: 2015.

Study-design: Retrospective chart review and analysis of clinical experiences.

Impact Factor: 1.839 (2015).

Patient collective: 80 adult patients (97 cases) treated between June 2012 and September 2013. Two-thirds of the patients had a BMI > 25 kg/m². 84% were classified as ASA I and II.

Methods: Patients underwent HFJV using thin, subglottically placed catheters. Analysis focused on complications and practical steps. The mean operating time averaged 53 minutes, and the mean duration of anesthesia was 81 minutes.

**Outcome: The aim was to evaluate the use and safety of HFJV in laryngotracheal surgery. Two reversible desaturations to 70%, one emergency re-intubation due to saturation drop below 50%, and eight elective re-intubations due to suboptimal oxygenation.**

Quality Score: 4

#### 3.1.26 In-circuit high-frequency jet ventilation to reduce organ motion in a child undergoing sarcoma ablation

Elgie LD, McPherson K, Yeung J, Marshall L, Windsor R, Bandula S.

Year: 2021.

Study-design: Case report.

Impact Factor: 0.64.

Patient collective: 7-year-old girl, 23 kg, with rare alveolar soft part sarcoma of the nasopharynx, requiring CT-guided microwave ablation of a metastatic right lung nodule under general anesthesia.

Methods: General anesthesia was induced and maintained with propofol and remifentanil. After intubation, HFJV was used for 45 minutes to identify and ablate the nodule. A jet swivel adapter was used to ensure optimal ventilation.

**Outcome: The ablation was successfully performed without complications. The patient recovered well and was discharged the following day. CT scans showed successful tumor ablation with no new disease or complications.**

Quality Score: 4

#### 3.1.27 Efforts to Enhance Catheter Stability Improve Atrial Fibrillation Ablation Outcome

Hutchinson MD, Garcia FC, Mandel JE, Elkassabany N, Zado ES, Riley MP, Cooper JM, Bala R, Frankel DS, Lin D, Supple GE, Dixit S, Gerstenfeld EP, Callans DJ, Marchlinski FE.

Year: 2013.

Study-design: Observational cohort study.

Impact Factor: 4.866 (2013).

Patient collective: 300 patients undergoing AF ablation, divided into 3 equal groups of 100 patients each. Group 1: AF ablation without I-EAM, SI, or HFJV. Group 2: AF ablation with I-EAM and SI, but without HFJV. Group 3: AF ablation with I-EAM, SI, and HFJV. Exclusion criteria included prior AF ablation, enrollment in clinical trials using investigational ablation systems, and procedures not performed according to prescribed study interventions.

Methods: 300 patients undergoing AF ablation were divided into 3 groups based on the tools utilized. Group 1 had no advanced tools, Group 2 used I-EAM and SI, and Group 3 used I-EAM, SI, and HFJV. The primary outcome was freedom from AF 1 year after a single ablation procedure. Acute and chronic pulmonary vein reconnection was also assessed.

**Outcome: This study examined the impact of modern stabilization techniques on the outcome of AF ablation. The incorporation of contemporary tools significantly improved 1-year freedom from AF after ablation (52% vs 66% vs 74%, P = .006) and reduced acute and chronic pulmonary vein reconnections.**

Quality Score: 3

#### 3.1.28 High-frequency jet ventilation during video-assisted thoracoscopic surgery in a patient with previous contralateral pneumonectomy

Ishiyama T, Iwashita H, Shibuya K, Terada Y, Masamune T, Nakadate Y, Matsukawa T.

Year: 2013.

Study-design: Case report.

Impact Factor: 1.016 (2012).

Patient collective: 77-year-old male, 45 kg, 160 cm, with right pneumothorax and previous left pneumonectomy.

Methods: HFJV was applied during VATS. Anesthesia was induced and maintained with propofol and fentanyl. A single-lumen endotracheal tube was used. HFJV was set at a frequency of 180 breaths/min, I/E ratio of 0.5, driving pressure of 0.5–0.8 bar, and FiO2 of 0.6 to 1.0.

**Outcome: PaO2 was well preserved, but PaCO2 gradually increased from 51.9 mmHg to 80.0 mmHg. Blood pressure and heart rate remained stable. Postoperative hypercapnia required continued positive pressure ventilation.**

Quality Score: 4

#### 3.1.29 Frequency dependence of lung volume changes during superimposed high-frequency jet ventilation and high-frequency jet ventilation

Sütterlin R, Priori R, Larsson A, Lo Mauro A, Frykholm P, Aliverti A.

Year: 2014.

Study-design: Single cohort study with a crossover design.

Impact Factor: 3.749 (2014).

Patient collective: Animals: 10 pigs (age approximately 3 months, weight 23–27 kg). Premedication: Xylazine (2.2 mg/kg), tiletamine (3 mg/kg), zolazepam (3 mg/kg). Intubation and tracheotomy: After bolus injection of fentanyl (100–500 µg intravenously). Anesthesia: Pentobarbital (7–9 mg/kg/h) and morphine (420–540 µg/kg/h). Muscle relaxant: Pancuronium (280–360 µg/kg/h).

Methods: SHFJV or HFJV were used alternately to ventilate the lungs of pigs. The low-frequency component was kept at 16 min−1 in SHFJV. In both modes, high frequencies ranging from 100 to 1000 min−1 were applied in random order, and ventilation was maintained for 5 minutes in all modalities. Chest wall volume variations were obtained using opto-electronic plethysmography. Airway pressures and arterial blood gases were measured repeatedly.

**Outcome: SHFJV was more effective in increasing end-expiratory volume compared to HFJV, but both modes provided adequate ventilation except HFJV at frequencies ≥300 min−1. SHFJV enabled higher, frequency-independent oxygenation and CO2 elimination.**

Quality Score: 3

#### 3.1.30 Reproducibility of target coverage in stereotactic spot scanning proton lung irradiation under high frequency jet ventilation

Santiago A, Jelen U, Ammazzalorso F, Engenhart-Cabillic R, Fritz P, Mühlnickel W, Enghardt W, Baumann M, Wittig A.

Year: 2013.

Study-design: Non-randomized controlled cohort.

Impact Factor: 5.060 (2013).

Patient collective: Patients with 12 lesions, including 9 with peripheral stage I NSCLC and 2 with metastases. Inclusion criteria: Target motion amplitude exceeding 1 cm.

Methods: Datasets of patients treated with single-fraction photon stereotactic radiosurgery under HFJV were analyzed. Scanned-beam proton plans were prepared using the TRiP98 treatment planning system with 2, 3–4, and 5–7 beams. The planning objective was to deliver at least 95% of the prescription of 33 Gy (RBE) to 98% of the PTV. Plans were subsequently recomputed on localization CT scans. Effects of range uncertainties were investigated for selected cases. Treatment involved single-fraction stereotactic radiotherapy with up to 33 Gy at the isocenter under HFJV.

**Outcome: The study investigates the reproducibility of target coverage in stereotactic spot scanning proton lung irradiation under HFJV. Median GTV V98% was 98.7% in the original 2-field plans and 93.7% in their recomputation (p=0.039). The respective values were 99.0% and 98.0% (p=0.039) for the 3–4-field plans and 100.0% and 99.6% (p=0.125) for the 5–7-field plans. HFJV provided reproducible tumor fixation, ensuring excellent target coverage in most cases, although setup errors affected dosimetric results in a few cases.**

Quality Score: 3

#### 3.1.31 Flow dynamics using high-frequency jet ventilation in a model of bronchopleural fistula

Wood MJ, Lin ES, Thompson JP.

Year: 2013.

Study-design: Non-randomized, controlled laboratory experiment.

Impact Factor: 9.8 (2013).

Patient collective: Cadaveric porcine lungs with a simulated bronchopleural fistula, artificial test lungs. The left pig lung was prepared and placed into an airtight acrylic box for experiments.

Methods: A 2 or 10 mm length fistula was created at proximal, middle, or distal sites in standard artificial ventilator “test” lungs and cadaveric porcine lungs. The effects of alterations in frequency, applied pressure, and entrained, expired, and leak volumes were determined using gauge and differential pressure sensors.

**Outcome: The study found that higher jet frequencies (>200 min⁻¹) and lower driving pressures (≤1.5 bar) may reduce air leaks and maintain ventilator volumes. The leak volume increased with larger and more proximally situated fistulas.**

Quality Score: 3

#### 3.1.32 Efficacy of target-controlled infusion of propofol and remifentanil with high frequency jet ventilation in fibre-optic bronchoscopy

Wang H, Yang C, Zhang B, Xia Y, Liu H, Liang H.

Year: 2013.

Study-design: Randomized controlled trial.

Impact Factor: 1.0.

Patient collective: 92 patients aged 20–75 years, scheduled for flexible bronchoscopy. Inclusion criteria: Patients aged 20–75 years undergoing flexible bronchoscopy. Exclusions: FEV1 < 1.0 L, ASA status > Class III, BMI > 25 kg/m², moderate/severe kidney or liver impairment, hypoxaemia needing supplementary oxygen at rest.

Methods: 92 patients were randomly assigned to receive either MS using TCI-delivered propofol and remifentanil (n=46) or GA using TCI-delivered propofol and remifentanil with HFJV (n=46). The study compared the incidence of hypoxaemia, cough score, haemodynamic parameters, duration of bronchoscopy, and patient satisfaction between groups.

**Outcome: This study evaluates the efficacy and safety of target-controlled infusion of propofol and remifentanil combined with HFJV to achieve general anesthesia in diagnostic flexible bronchoscopy. GA with HFJV showed a lower incidence of hypoxaemia (2.2% vs. 34.1%), lower cough score, shorter bronchoscopy duration, and higher patient satisfaction compared to MS.**

Quality Score: 2

#### 3.1.33 High-frequency jet ventilation shortened the duration of gas embolization during laparoscopic liver resection in a porcine model

Fors D, Eiriksson K, Waage A, Arvidsson D, Rubertsson S.

Year: 2014.

Study-design: Randomized controlled trial.

Impact Factor: 5.616 (2014).

Patient collective: 24 Swedish country-bred piglets, aged approximately 3 months, of both sexes, fasted overnight with free access to water. Piglets were randomized into two groups (12 per group): normal frequency ventilation (NFV) and HFJV.

Methods: Piglets were randomized to NFV or HFJV groups. Liver lobe resection (LLR) with a standardized injury to the left hepatic vein was performed. Hemodynamic and respiratory variables were monitored. Transesophageal echocardiography was used to detect and grade gas embolism (GE). Grading of GE: Grade 0 (<5 bubbles), Grade 1 (≥5 bubbles but not completely obscuring the outflow tract), Grade 2 (completely obscuring the outflow tract).

**Outcome: The study aimed to explore whether HFJV could reduce the frequency, severity, or duration of GE during LLR. Results indicated that the mean duration of GE was shorter in the HFJV group (P=0.008). However, there were no significant differences between the groups in terms of GE frequency or severity. The physiological responses to GE were variable and unpredictable. HFJV was found to be a feasible ventilation method during LLR and may help in minimizing the duration of GE without affecting the frequency or severity of embolic episodes.**

Quality Score: 2

#### 3.1.34 High-frequency jet ventilation under general anesthesia facilitates CT-guided lung tumor thermal ablation compared with normal respiration under conscious analgesic sedation

Chung DYF, Tse DML, Boardman P, Gleeson FV, Little MW, Scott SH, Anderson EM.

Year: 2014.

Study-design: Retrospective cohort study.

Impact Factor: 2.8.

Patient collective: 26 patients in the HFJV group vs. 13 in the normal respiration (NR) group. Gender: 16 men and 10 women (HFJV) vs. 8 men and 5 women (NR). Age: 66.39 ± 12.20 years (HFJV) vs. 69.15 ± 15.59 years (NR). Weight: 80.52 ± 19.75 kg (HFJV) vs. 74.54 ± 22.63 kg (NR). Body mass index (BMI): 27.80 ± 5.73 kg/m² (HFJV) vs. 26.15 ± 6.84 kg/m² (NR). ASA classification: 2.66 ± 0.48 (HFJV) vs. 2.23 ± 0.44 (NR). FEV1: 2.08 ± 0.90 (HFJV) vs. 2.32 ± 0.75 (NR). Previous ipsilateral hemithoracic interventions: 10 (HFJV) vs. 4 (NR). Tumor characteristics: 47 tumors in the HFJV group vs. 16 in the NR group. Tumor size: 16.09 ± 9.21 mm (HFJV) vs. 27.38 ± 14.73 mm (NR).

Methods: Thermal ablation treatment sessions were reviewed for both HFJV under general anesthesia (GA) and normal respiration (NR) under conscious sedation (CS). Measures of technical difficulty, including time duration, number of CT fluoroscopic acquisitions, and radiation dose, were compared. Anesthesia and procedural records were analyzed, and statistical comparisons were made using Student’s independent-samples t-test and x² test.

**Outcome: Patients treated with HFJV under GA had higher ASA classifications and smaller tumors. The time required for applicator placement, number of CT shots, and radiation dose were significantly lower with HFJV under GA. There were no significant differences in anesthesia time or complication rates.**

Quality Score: 3

#### 3.1.35 High-frequency jet ventilation using the Arndt bronchial blocker for refractory hypoxemia during one-lung ventilation in a myasthenic patient with asthma

El-Tahan MR, Doyle DJ, Hassieb AG.

Year: 2014.

Study-design: Case series.

Impact Factor: 1.006.

Patient collective: 66-year-old patient, 162.5 cm, 97.5 kg. Medical history: Myasthenia gravis (Osserman class IIA), moderate bronchial asthma, diabetes mellitus, hiatal hernia, cervical disc herniation. Medications: Pyridostigmine (360 mg/day), prednisolone (30 mg/day), insulin, metformin, inhaled steroid, ipratropium, salbutamol.

Methods: During the surgery, one-lung ventilation (OLV) was performed using an Arndt bronchial blocker. Due to severe, refractory hypoxemia, high frequency jet ventilation (HFJV) was applied through the Arndt bronchial blocker. This approach aimed to improve oxygenation and optimize ventilation parameters.

Outcome: The paper presents a case where HFJV through an Arndt bronchial blocker was successfully used to manage severe hypoxemia during OLV. There was significant improvement in oxygenation, reduced airway pressures, and effective treatment of refractory hypoxemia during OLV.

Quality Score: 4

#### 3.1.36 High-Frequency Jet Ventilation against Small-Volume Conventional Mechanical Ventilation in the Rabbit Models of Neonatal Acute Lung Injury

Mokra D, Tomcikova Mikusiakova L, Mikolka P, Kosutova P, Jurcek M, Kolomaznik M, Calkovska A.

Year: 2016.

Study-design: Experimental animal study. Impact Factor: 2.0.

Patient collective: 2 adult New Zealand white rabbits of both genders with a mean body weight of 2.5 ± 0.3 kg.

Methods: Rabbits were ventilated with HFJV or CMV for 4 hours after inducing lung injury by saline lung lavage (LAV) or meconium aspiration syndrome (MAS). Ventilatory pressures, blood gases, and indexes of gas exchange were assessed.

**Outcome: The study evaluated the potential usefulness of HFJV compared to CMV in acute lung injury. HFJV was less effective than CMV at 1 hour, but after 4 hours, both methods showed comparable lung function and edema formation.**

Quality Score: 4

#### 3.1.37 Technical description of a modified jet ventilation injector for airway laser surgery in neonates and infants: retrospective analysis of 20 cases

Martins MR, Van Boven M, Schmitz S, Hamoir M, Veyckemans F.

Year: 2016.

Study-design: Retrospective analysis.

Impact Factor: 2.245 (2016).

Patient collective: 20 cases of neonates and infants (5 boys, 4 girls) with a mean age of 7.4 ± 6.9 months and a mean weight of 6 ± 2.8 kg. A total of 20 procedures were performed. Pathologies included vocal cord granulomas, laryngeal papillomatosis, vocal cord papillomas, saccular cysts, laryngomalacia, vallecular tongue cysts, and thyroglossal cysts.

Methods: Retrospective review of the anesthetic records of all children younger than 2 years undergoing transglottal HFJV for CO₂ laser laryngeal procedures using a modified adult injector between 2006 and 2013.

**Outcome: The authors modified an adult jet ventilation injector to provide transglottal high frequency jet ventilation in small children undergoing CO₂ laser laryngeal procedures. No complications were observed with the use of HFJV or the modified injector. The modified device ensured excellent visibility and field access for the surgeon as well as adequate ventilation during surgery.**

Quality Score: 4

#### 3.1.38 Assessing Initial Response to High-Frequency Jet Ventilation in Premature Infants with Hypercapnic Respiratory Failure

Wheeler CR, Smallwood CD, O’Donnell I, Gagner D, Sola-Visner MC.

Year: 2017.

Study-design: Retrospective single-center study.

Impact Factor: 2.066.

Patient collective: 34 preterm infants (24 males), transferred to the neonatal ICU at Boston Children’s Hospital for further medical or surgical management. Inclusion criteria: Birth weight < 2,000 g and capillary pCO2 > 55 mm Hg. Exclusion criteria: Non-repaired complex congenital heart defects, birth weight > 2,000 g, HFJV at the time of admission. Study period: January 2012 to January 2016.

Methods: Ventilator parameters and physiological data were recorded 1 hour before and at 1, 4, and 6 hours after the initiation of HFJV. Subjects were classified as responders if their capillary pCO2 decreased by more than 10% after 1 hour of HFJV.

**Outcome: The study examined the effectiveness of HFJV in preterm infants with hypercapnic respiratory failure. Of the 34 premature infants studied, 25 responded with >10% reduction in capillary pCO2 after 1 hour. Responders showed improved pH and reduced FIO2 within the first hour. Non-responders had higher initial peak inspiratory pressures and were older.**

Quality Score: 3

#### 3.1.39 Kasuistik: Suffiziente High-Frequency-Jet-Ventilation über 2,5 h

Kampe S, Rocha M, Darwiche K, Ebmeyer U, Georgios S.

Year: 2016.

Study-design: Case series.

Impact Factor: 0.273.

Patient collective: 54-year-old patient, 162 cm, 44 kg, ASA classification III. Medical history includes Ross procedure (2009), aortic arch replacement (2013), aortic root replacement with valve-carrying conduit (Bentall, 23 mm bioprosthesis), and pulmonary valve replacement with homograft. Current conditions include functional narrowing of the trachea at the bifurcation (50%) and a persistent esophagotracheal fistula (∼2 cm).

Methods: HFJV was applied using a jet catheter with 1.5 mbar pressure and a frequency of 150/min. Total intravenous anesthesia was administered with propofol and remifentanil. Rigid bronchoscopy and tracheal stent removal were performed, with HFJV maintained for 2.5 hours during surgery.

**Outcome: The patient underwent successful surgery to close the persistent esophagotracheal fistula and perform a tracheal segment resection. Postoperatively, the patient exhibited stable cardiopulmonary values and acid-base balance. The recovery was uncomplicated, and the patient was discharged on the 17th day.**

Quality Score: 4

#### 3.1.40 Elective use of the Ventrain for upper airway obstruction during high-frequency jet ventilation

Fearnley RA, Badiger S, Oakley RJ, Ahmad I.

Year: 2016.

Study-design: Case series.

Impact Factor: 2.163 (2020).

Patient collective: 58-year-old male, 56 kg, 172 cm, with a history of squamous cell carcinoma of the right vocal cord and anterior commissure.

Methods: Elective use of Ventrain for ventilation in a patient with upper airway obstruction. Fiber optic intubation was performed under topical anesthesia and sedation. Ventilation was achieved using the Ventrain device with an I:E ratio of 1:1 at a rate of 10–12 breaths per minute, driven by 15 L/min oxygen.

**Outcome: The article highlights the successful elective use of the Ventrain ventilation device during HFJV in a patient with upper airway obstruction. Ventilation and surgery were successfully maintained for over 60 minutes, with oxygen saturations at 100% and ETCO₂ levels between 5.4 and 6.0 kPa. No major complications occurred post-surgery.**

Quality Score: 4

#### 3.1.41 Recovery and safety with prolonged high-frequency jet ventilation for catheter ablation of atrial fibrillation: A hospital registry study from a New England healthcare network

Munoz-Acuna R, Tartler TM, Azizi BA, Suleiman A, Ahrens E, Wachtendorf LJ, Linhardt FC, Chen G, Tung P, Waks JW, Schaefer MS, Sehgal S.

Year: 2024.

Study-design: Case series.

Impact Factor: 5.0 (2023).

Patient collective: The study’s patient cohort consisted of 1,822 individuals who underwent catheter ablation for atrial fibrillation. Patients were aged 18 years or older, with a mean age of 62 years (±10 years), and 29% of the participants were female. Eligibility for inclusion required the availability of electronic documentation of exposure and primary outcome variables.

Of the 1,822 patients, 1,157 (63%) received high-frequency jet ventilation (HFJV), while 665 (37%) were managed with conventional ventilation. Patients requiring emergency procedures or those with an ASA (American Society of Anesthesiologists) status of V or higher were excluded from the study.

Methods: The primary outcome evaluated was the length of stay in the post-anesthesia care unit (PACU). Secondary analyses examined the impact of high-frequency jet ventilation (HFJV) on intra-procedural hypoxemia, defined as peripheral hemoglobin oxygen saturation (SpO2) below 90%. Additionally, post-procedural respiratory complications (PRC), intra-procedural hypocarbia, and hypotension were assessed. The analyses utilized multivariable negative binomial and logistic regression models, adjusted for patient and procedural characteristics.

**Outcome: The median length of stay in the post-anaesthesia care unit (PACU) was 226 minutes (IQR: 163–361 minutes) for patients receiving HFJV, compared to 244 minutes for those with conventional ventilation. However, after adjustment, HFJV was associated with a significantly longer PACU stay (adjusted difference: 37.7 minutes; 95% confidence interval [CI]: 9.7–65.8; p = 0.008).**

**Patients undergoing HFJV had a higher risk of intra-procedural hypocapnia (adjusted odds ratio [ORadj]: 5.90; 95% CI: 2.63–13.23; p < 0.001) and hypotension (ORadj: 1.88; 95% CI: 1.31–2.72; p = 0.001). No significant association was found between HFJV and intra-procedural hypoxaemia (p = 0.51) or postoperative respiratory complications (PRC; p = 0.97).**

Quality Score: 4

### 3.2 Comparison of HFJV based on safety and performance parameters

**Table 4:**
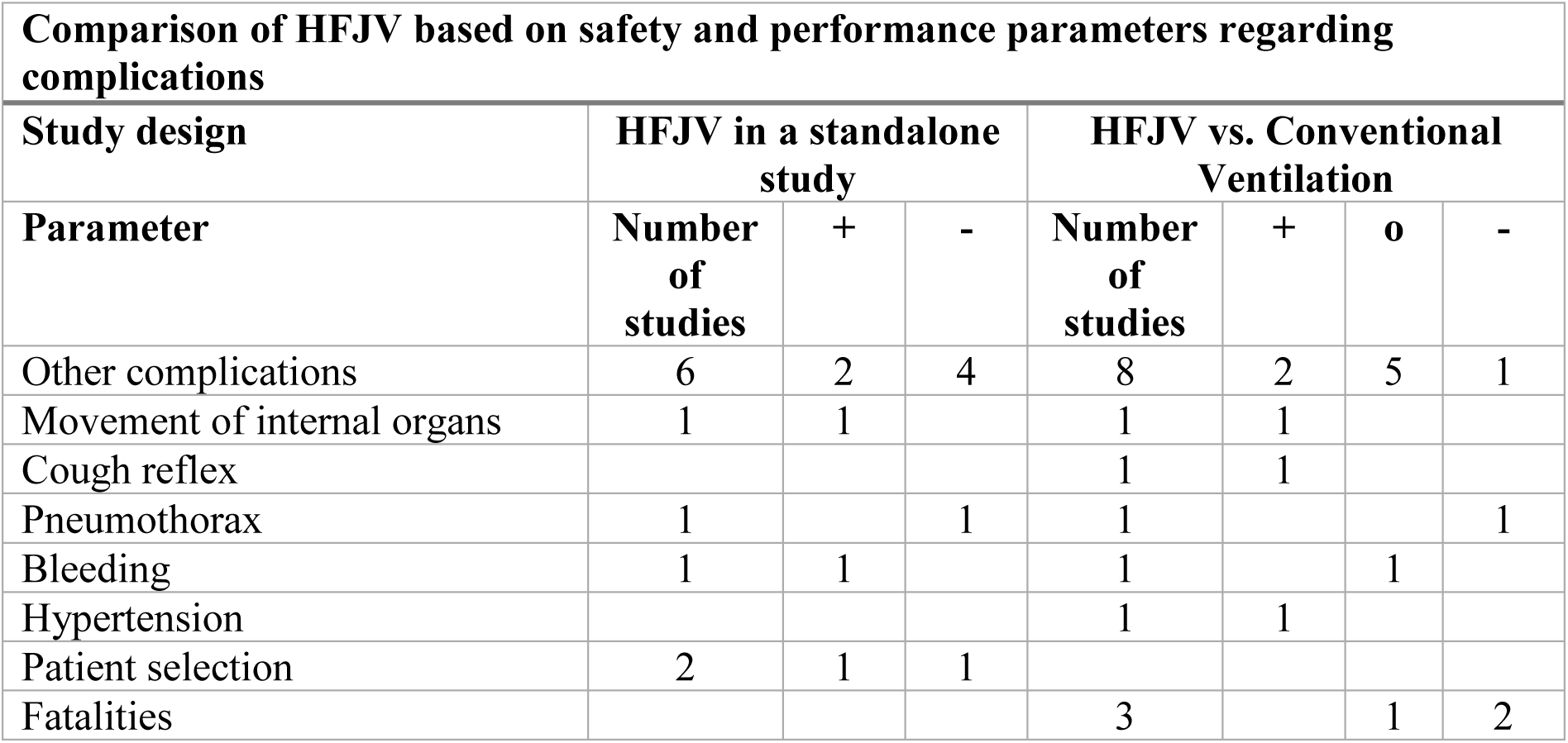
Results Table 1 – Complications.

**Table 5:**
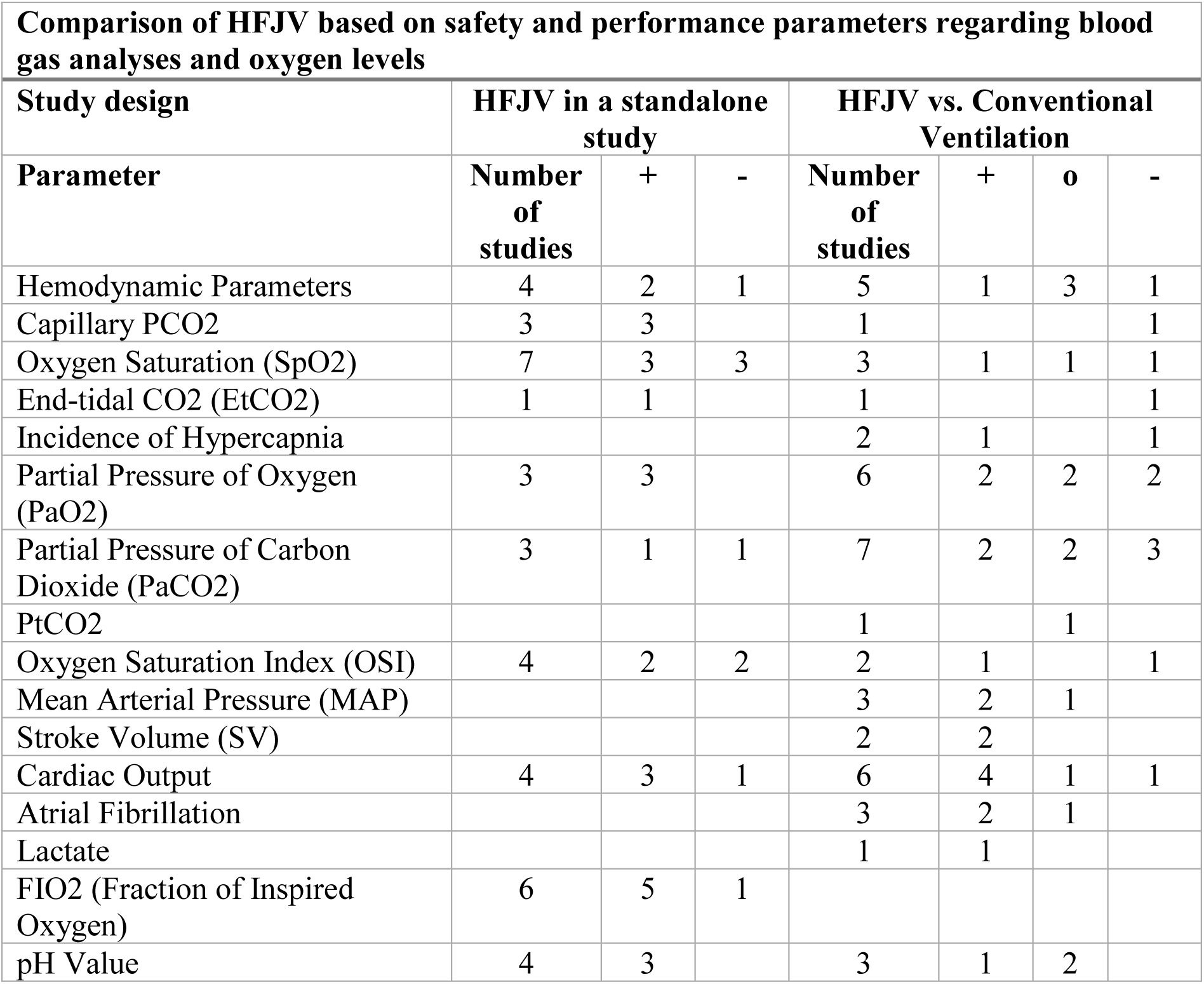
Results Table 2 - Blood Analysis and Oxygen Values.

**Table 6:** Results Table 3 - Ventilation Parameters.

| Comparison of HFJV based on safety and performance parameters regarding ventilation parameters |  |  |  |  |  |  |  |
| --- | --- | --- | --- | --- | --- | --- | --- |
| Study design | HFJV in a standalone study |  |  | HFJV vs. Conventional Ventilation |  |  |  |
| Parameter | Number of studies | + | - | Number of studies | + | o | - |
| PIP: Peak Inspiratory Pressure | 1 | 1 |  | 1 | 1 |  |  |
| PEEP: Positive End-Expiratory Pressure | 2 |  |  | 1 |  |  | 1 |
| Tidalvolumen: Tidal Volume | 1 | 1 |  | 3 | 1 |  | 2 |
| Airway Pressure (Paw) | 4 | 3 |  | 1 | 1 |  |  |
| Pressure above PEEP | 1 | 1 |  | 1 |  |  | 1 |
| Inspiratory Time |  |  |  | 1 |  |  | 1 |
| Impedance Reduction |  |  |  | 1 | 1 |  |  |
| Kontaktkraft: Contact Force |  |  |  | 2 | 2 |  |  |

**Table 7:** Results Table 4 – Treatments.

| Comparison of HFJV based on safety and performance parameters regarding treatments |  |  |  |  |  |  |  |
| --- | --- | --- | --- | --- | --- | --- | --- |
| Study design | HFJV in a standalone study |  |  | HFJV vs. Conventional Ventilation |  |  |  |
| Parameter | Number of studies | + | - | Number of studies | + | o | - |
| Treatment duration | 2 | 2 |  | 7 | 3 | 1 | 3 |
| Stable ventilation | 2 | 2 |  |  |  |  |  |
| Treatment process | 8 | 8 |  | 4 | 4 |  |  |

**Table 8:** Results Table 5 - Follow-up Treatments and Postoperative Parameters.

| Comparison of HFJV based on safety and performance parameters regarding post-treatment and postoperative parameters |  |  |  |  |  |  |  |
| --- | --- | --- | --- | --- | --- | --- | --- |
| Study design | HFJV in a standalone study |  |  | HFJV vs. Conventional Ventilation |  |  |  |
| Parameter | Number of studies | + | - | Number of studies | + | o | - |
| Follow-up treatments |  |  |  | 1 | 1 |  |  |
| Postoperative treatments |  |  |  | 2 |  |  | 2 |

**Table 9:**
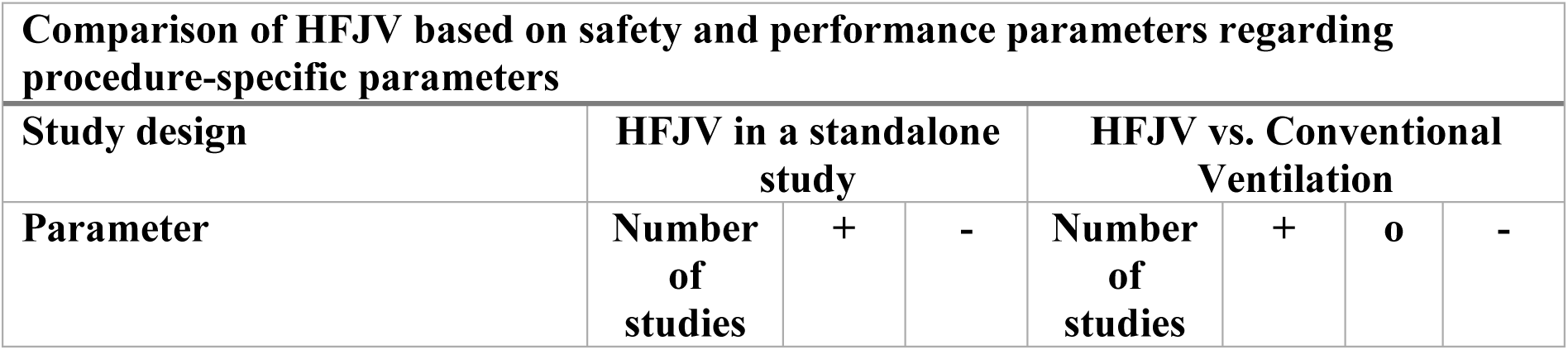
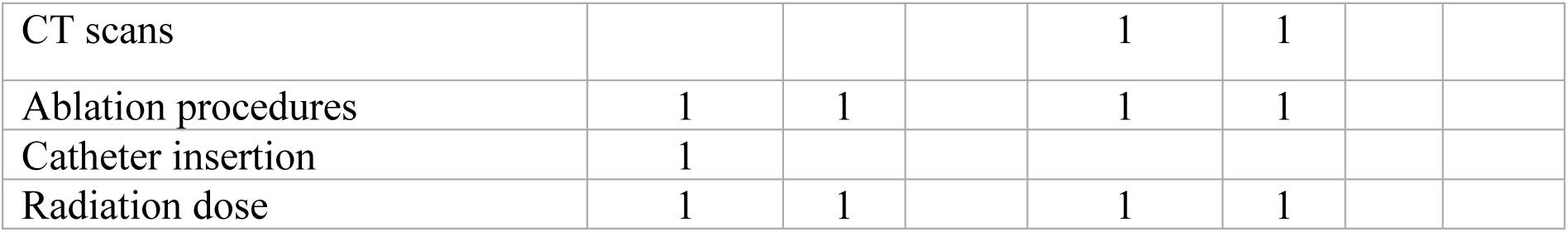
Results Table 6 - Procedure-Specific Parameters.

| Comparison of HFJV based on safety and performance parameters regarding procedure-specific parameters |  |  |  |  |  |  |  |
| --- | --- | --- | --- | --- | --- | --- | --- |
| Study design | HFJV in a standalone study |  |  | HFJV vs. Conventional Ventilation |  |  |  |
| Parameter | Number of studies | + | - | Number of studies | + | o | - |
| CT scans |  |  |  | 1 | 1 |  |  |
| Ablation procedures | 1 | 1 |  | 1 | 1 |  |  |
| Catheter insertion | 1 |  |  |  |  |  |  |
| Radiation dose | 1 | 1 |  | 1 | 1 |  |  |

**Table 10:** Results Table 7 - Usability and Patient Satisfaction.

| Comparison of HFJV based on safety and performance parameters regarding usability and patient satisfaction |  |  |  |  |  |  |  |
| --- | --- | --- | --- | --- | --- | --- | --- |
| Study design | HFJV in a standalone study |  |  | HFJV vs. Conventional Ventilation |  |  |  |
| Parameter | Number of studies | + | - | Number of studies | + | o | - |
| Anesthesiological monitoring | 1 |  | 1 |  |  |  |  |
| Easy handling | 1 | 1 |  |  |  |  |  |
| Mechanical problems | 1 |  | 1 | 2 | 1 |  | 1 |
| Patient satisfaction |  |  |  | 1 | 1 |  |  |

**Legend:**

**Comparison of HFJV based on safety and performance parameters:**

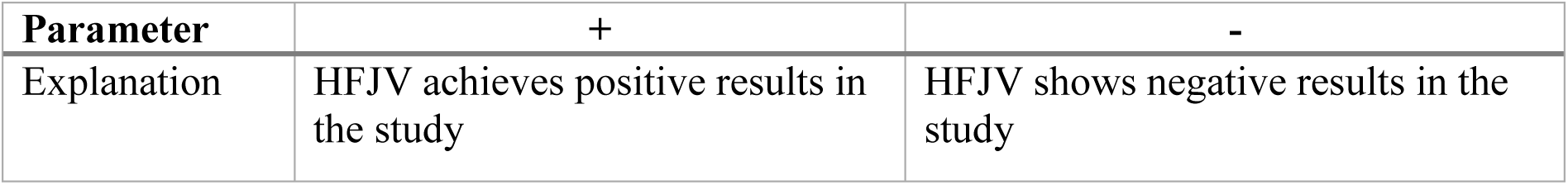

| Parameter | + | - |
| --- | --- | --- |
| Explanation | HFJV achieves positive results in the study | HFJV shows negative results in the study |

**HFJV compared to conventional ventilation based on safety and performance parameters:**

| Parameter | + | o | - |
| --- | --- | --- | --- |
| Explanation | HFJV is more effective compared to another ventilation model | No significant difference between HFJV and another ventilation model | HFJV is less effective compared to another ventilation model |

In the seven result tables, the safety and performance parameters of high frequency jet ventilation were analysed. For this investigation, 40 studies were selected and systematically categorised according to safety and performance criteria.

The findings were divided into two tables. The first table includes all studies in which HFJV was examined as a standalone ventilation method, while the second table contains studies comparing HFJV with conventional ventilation. Studies showing positive outcomes (“+”) indicate that HFJV offers advantages in terms of the assessed safety and performance parameters, demonstrating its efficacy and benefit. In contrast, studies with negative outcomes (“–”) highlight potential limitations or risks associated with HFJV.

In direct comparison with conventional ventilation, the evaluated studies were grouped into three categories. Studies with positive outcomes (“+”) illustrate that HFJV is more effective and safer than conventional ventilation methods. Studies showing no significant difference (“o”) indicate comparable safety and performance parameters between HFJV and conventional ventilation. Studies with negative outcomes (“–”) suggest that HFJV may be less effective or pose higher risks in specific clinical situations as compared to conventional ventilation.

Most studies reveal no significant differences in complication rates between HFJV and conventional ventilation (Table 1). From 20 studies, two reported negative outcomes and four noted increased complications with exclusive use of HFJV. In two studies, pneumothorax occurred, mainly during prolonged procedures, and one study documented pulmonary bleeding during HFJV.

Additional complications were described in several studies, with five finding no significant differences from conventional ventilation, while five reported adverse events. Two of the 40 studies noted fatalities following HFJV, although one of these reported improvements compared to conventional ventilation. Challenges with overweight patients were also mentioned, but no differences in overall mortality were observed.

HFJV demonstrated positive effects in reducing organ movement and radiation dose (Table 4), with six studies reporting these benefits. Furthermore, pH and FiO₂ values remained stable (Table 2). Two out of 18 studies indicated worsened oxygenation, while ten found no significant changes. The partial pressure of carbon dioxide showed no differences or improvement in six out of ten studies. Two studies noted mechanical issues during HFJV, while others rated its handling as advantageous (Table 7).

## 4 Discussion

The application of high frequency jet ventilation across various clinical settings demonstrates both advantages and challenges. Numerous studies indicate the efficacy of HFJV, particularly in reducing organ motion and necessary radiation dose, as well as in managing patients with severe respiratory problems. However, the use of HFJV requires careful consideration of the specific risks and potential complications associated with this ventilation technique.

One of the most frequently documented complications of HFJV is pneumothorax, which can result from excessive ventilation pressure or mechanical damage. The usual mechanism of this complication is the inadvertent obstruction of the passive gas outflow, thus leading to rapid gas trapping. Prospective study No. 7 (Quality III) reports a pneumothorax rate of 1%, with some cases necessitating chest drainage [17]. Although the incidence is relatively low, the clinical significance of this complication highlights the need for precise pressure control. The non-randomised cohort study has methodological limitations but provides valuable insights into the risk profile of HFJV. A similar observation was made in study No. 20, which documented four cases of pneumothorax in neonates, underscoring the vulnerability of this patient group and the importance of careful pressure control [18]. However, the limited sample size, classification as Quality Level IV, and the selection of the most severe cases limit the generalisability of these findings.

In addition to pneumothorax, subcutaneous emphysema is another relevant complication, often occurring after multiple tracheal punctures. Study No. 7 reports an incidence rate of 8.4% for subcutaneous emphysema, caused by the escape of gas into the subcutaneous tissue, leading to swelling and a sensation of pressure [17]. This complication underscores the importance of optimal tracheal cannula placement and close monitoring during HFJV to minimise the risk of such adverse events.

Patients with severe COPD, obesity, or other pulmonary diseases are particularly susceptible to complications such as hypoventilation, hypercarbia by CO_2_ retention, and arterial desaturation under HFJV. Studies indicate that precise monitoring of ventilation parameters in all patient is essential to avoid adverse events [17, 19, 20].

Inadequate peak pressure limitation and errors in airway management can increase the risk of serious complications, such as pneumothorax, barotrauma, and subcutaneous emphysema, which is why continuous monitoring and precise adjustment of ventilation parameters are essential [17, 18, 21, 22].

A similar observation was made in retrospective case series No. 41 (Quality IV) from 2024, which investigated the effects of HFJV in 1,822 patients undergoing catheter ablation for atrial fibrillation. The findings revealed that while HFJV improved intraoperative catheter stability, it was associated with an increased risk of intraoperative hypocapnia and hypotension [23].

Of particular concern is the observation that the prolonged recovery time in the post-anaesthesia care unit for patients undergoing HFJV might be attributed to insufficient monitoring and adjustment of ventilation parameters. This study highlights the role of higher working pressures and the resulting hypocapnia, emphasising the necessity of continuous monitoring of pCO₂ levels during the procedure. Additionally, differences in anaesthetic approaches, particularly the use of propofol and remifentanil in the HFJV group compared to volatile agents in the control group, may represent a significant confounding factor.

The findings underline the importance of standardised protocols for ventilation management and anaesthesia monitoring to minimise the potential risks associated with HFJV and ensure patient safety. Particularly crucial is the precise adjustment of ventilation settings, such as working pressure and respiratory rate, alongside continuous monitoring of pCO₂ levels. This approach would help mitigate complications like hypocapnia and hypotension [23, 24].

The use of HFJV requires comprehensive knowledge and experience on the part of the involved staff. Problems mainly occur in units, where HFJV is only rarely used, and the staff is unable to acquire sufficient experience. Precise control of ventilation parameters is particularly crucial in complex procedures and high-risk patients to avoid potential complications [17, 21]. This is especially relevant for preterm infants and neonates, as retrospective studies like No. 19 (Quality IV) demonstrate a significantly higher mortality rate in the HFJV group (24%) compared to the control group (4%) among critically ill preterm infants [25]. These findings highlight that pneumothorax and other complications can have serious consequences, and that HFJV in this vulnerable patient group should be approached with utmost caution.

Nevertheless, several studies demonstrate positive effects of HFJV. Prospective study No. 38 (Quality II) examined the use of HFJV in preterm infants with hypercapnic respiratory failure and found significant improvements in blood gas levels and enhanced CO₂ elimination, underscoring the importance of HFJV for neonatal patients [26]. Study No. 16 (Quality III) also reported reduction in PaCO₂ and the oxygenation index within the first 24 hours of ventilation in preterm infants, albeit with limitations due to small sample size and lack of randomisation [27].

Studies No. 37 and No. 5 support the safe use of high frequency jet ventilation in neonatal patients. Retrospective analysis No. 37 (Quality IV) documented HFJV use in neonates and infants, particularly with modified injectors for transglottal HFJV. In this report of 20 cases, no complications were observed, emphasising the suitability of the modified injector for transglottal application of HFJV [28].

In the non-randomised controlled cohort study No. 5, the use of HFJV in neonatal acute lung injury was examined. The findings indicated that HFJV could be safely applied while maintaining stable haemodynamic conditions. A key finding was the necessary increase in FiO₂ to 1.0 to improve oxygen saturation. Throughout the study, blood pH remained stable, indicating effective CO₂ elimination [29].

Although case study No. 37 and cohort study No. 5 are classified as being of low-quality evidence due to their design, they nonetheless provide valuable insights into the application and adjustment of HFJV in critical neonatal scenarios [28, 29].

There are also several studies in adult medicine supporting the use of HFJV. The randomised prospective study No. 18 (Quality II) compared superimposed high frequency jet ventilation (SHFJV) with conventional HFJV in terms of safety and effectiveness during bronchoscopic procedures. During the intervention, the HFJV group showed higher PaO₂ values compared to the SHFJV group, though this came with a higher risk of hypercapnia. Conversely, the SHFJV group demonstrated more stable PaCO₂ levels and a reduced incidence of hypercapnia, particularly during longer procedures. The study’s methodology is robust, as patient characteristics and comorbidities were balanced across the groups, enhancing the internal validity of the findings [30].

One major advantage of HFJV lies in its improved oxygenation and CO₂ elimination, especially in patients with refractory hypoxaemia and complex pulmonary interventions. Case series No. 35 on HFJV application in an adult patient with severe hypoxaemia demonstrated positive effects in this regard. The FiO₂ was raised to 1.0 to maximise oxygen delivery, and the stable pH indicated effective ventilatory efficiency. However, the quality of evidence in this study is lower, limiting the strength of its findings [31]. Additional positive outcomes were documented in studies No. 39 and No. 40. Case series No. 40 (Quality IV) describes the elective use of the Ventrain ventilation device to manage upper airway obstruction in a patient with post-radiation fibrosis during HFJV. Despite challenging airway conditions, stable respiratory parameters were maintained [32].

In Case study No. 39, the use of HFJV in complex airway surgical interventions was reported. HFJV was successfully maintained throughout the procedure, ensuring the patient’s acid-base balance and cardiopulmonary stability. No intraoperative complications occurred, and the postoperative course was uneventful [22].

Although case studies generally have low evidence quality, making their results less generalisable, they still offer valuable insights into the practical use of HFJV in patients with severe airway obstructions [22, 32].

The randomised controlled study No. 12 showed a significant improvement in catheter stability during atrial fibrillation ablation under HFJV. HFJV reduced the variability in contact force, resulting in more uniform ablation lesions, thereby increasing ablation success rates and shortening the duration of the intervention. The high quality and robust design of the study make the findings reliable and clinically relevant [33].

These results were further substantiated by studies No. 11 and No. 15. The randomised controlled study No. 11 (Quality II) examined the recurrence rate of atrial fibrillation in patients treated with either HFJV or standard ventilation. Results showed that the HFJV group had a significantly lower arrhythmia recurrence rate (31% vs. 50%, p = 0.019). Despite a higher use of vasopressors in the HFJV group, no significant additional complications were observed [34].

Similarly, study No. 15 (Quality III) offers valuable insights into HFJV use during atrial fibrillation ablation. Although this study is of lower quality than study No. 11, it also showed reduced arrhythmia recurrence rates without HFJV-specific complications, supporting HFJV application in cardiology, particularly for patients with atrial fibrillation [35].

HFJV demonstrates clear advantages in surgical oncology too. Several studies indicate that HFJV reduces ventilation dependent internal organ motion, particularly beneficial for minimally invasive and high-precision procedures. In the randomised controlled study No. 3 (Quality II), tumour ablations in the lung, liver, and kidney were investigated. HFJV achieved near-complete immobilisation of internal organs in 80% of patients, significantly enhancing needle placement precision and treatment success. The high quality of this randomised study strengthens the findings, especially for precise interventions in sensitive organs [19].

Another prospective, randomised, double-blind study (No. 9, Quality II) showed that HFJV significantly reduced intraoperative bleeding and improved visibility in the surgical field during functional endoscopic sinus surgery (FESS). These findings are particularly relevant, as clear visibility during surgery is crucial for success and complication minimisation. With an evidence grade of II, this study provides robust evidence that HFJV optimises the surgical field by lowering mean airway pressure and improving venous return [36].

Studies No. 21 (Quality III), No. 22 (Quality IV), case study No. 26 (Quality IV), and study No. 34 (Quality III) support the benefits of HFJV in minimising respiratory movements, especially advantageous for thoracic and abdominal surgeries. In study No. 21 (Quality III), HFJV was employed for tumour ablation, achieving near-complete internal organ immobilisation and significantly improving needle placement precision, with a successful ablation probe placement on the first attempt in 96% of cases. However, due to the single-cohort design without a control group, the quality of these findings is lower than that of randomised studies [37].

Case study No. 26 examined HFJV use in minimally invasive procedures, highlighting improved precision and safety. However, further research is needed, particularly to determine optimal parameters for paediatric patients [38].

Study No. 34 similarly found no specific HFJV-related incidents. General anaesthesia-related complications were similarly distributed across groups, supporting HFJV’s safety in clinical use [20].

HFJV also has potential to reduce procedural time and complication rates in certain surgeries. In the randomised controlled study No. 14 (Quality II), the duration of percutaneous dilatational tracheostomy was significantly shorter in the HFJV group than in the control group, with fewer complications like tube dislocations or desaturation events during the procedure [39].

Comparable advantages were noted in the randomised controlled study No. 32 (Quality II), which found that HFJV improved intraoperative visibility and control of organ movement during bronchoscopies, reducing procedural time [40].

Both studies, with evidence grade II, offer a solid foundation for evaluating the efficacy of HFJV in shortening operation duration and reducing complications [39, 40].

In the management of gas embolism, HFJV also shows benefits. Randomised controlled study No. 33 (Quality II) compared HFJV with conventional ventilation in laparoscopic procedures, examining the duration and severity of gas embolisms. Results indicated a significantly shorter embolism duration in the HFJV group, although there were no significant differences in frequency or severity between groups, suggesting HFJV as a potentially safer option. However, the study highlighted limitations, particularly concerning patients with severe respiratory diseases and obesity [18].

HFJV offers distinct advantages in specific clinical situations and should therefore be selectively employed. In neonates with hypercapnic respiratory failure, HFJV has proven effective, with significant improvements in blood gas values and more effective CO₂ elimination observed [27, 26].

In oncological surgery, particularly for tumour ablation in sensitive organs such as the lungs, liver, or kidneys, HFJV achieves near-complete organ immobilisation, greatly enhancing procedural precision and success rates [19]. HFJV is also beneficial in bronchoscopic procedures, providing more stable CO₂ levels and reducing hypercapnia incidence, especially during longer procedures [30]. Another area of application is atrial fibrillation ablation, where HFJV contributes to contact force stability and increases ablation success rates without significant additional complications [34, 33 ,35].

Despite HFJV’s advantages, there are patient groups for whom conventional ventilation may be preferable. For neonates with severe lung disease, several studies have reported a higher mortality rate with HFJV, highlighting this patient group’s particular vulnerability [25].

Additionally, patients with COPD, obesity, or other severe lung conditions should be ventilated conventionally, as HFJV increases the risk of pneumothorax and other complications in these cases [19, 20].

Precise control of ventilation parameters is essential to minimise risks such as pneumothorax, barotrauma, or subcutaneous emphysema. Accurate adjustment of ventilation pressures and continuous monitoring can help prevent these severe complications [17, 21]. For high-risk patients, such as those with COPD or in neonates, close monitoring is necessary to promptly detect and to counteract hypoventilation, arterial desaturation, and other ventilation complications [17, 25, 20].

High frequency jet ventilation imposes specific demands on conditioning the respiratory gas, which should always be considered regardless of the procedure duration. A common misconception is that heating and humidification are only necessary for longer interventions [41]. However, it has been shown that unconditioned gas during HFJV can quickly lead to airway mucosa dehydration and damage, significantly increasing the risk of complications [42].

The absence of gas conditioning may result in dehydration of the mucosa, which serves as a protective barrier and plays a vital role in lung hygiene [41]. Dry mucosa is more susceptible to injury, potentially leading to microfissures and crust formation. These changes increase the likelihood of inflammatory reactions and airway complications, particularly affecting the delicate structure of lung tissue [42].

Additionally, unconditioned air increases the risk of barotrauma, as cold and dry air reduces the elasticity of airway tissue. This may impair the flexibility and adaptability of the mucosa under pressure, elevating the risk of tissue injury. HFJV presents a unique strain on the airways due to the high speeds and pressures used to maintain gas exchange. Therefore, continuous conditioning is essential to protect mucosal integrity and tissue from thermal and mechanical stress [43, 41].

Furthermore, introducing cold, dry air without conditioning negatively impacts mucociliary clearance, a natural airway defense mechanism crucial for pathogen and foreign particle removal. Without sufficient humidification, mucociliary transport can be significantly slowed, increasing the risk of infectious complications and atelectasis due to secretion build-up [42,43]. In severe cases, this may lead to necrotising tracheobronchitis, a condition marked by intense inflammation and tissue necrosis within the airway [43]. The loss of ciliated epithelium and impaired mucociliary clearance due to inadequate conditioning further exacerbate mucus retention and can promote infectious complications [43].

Conditioning of the ventilatory gas is therefore a crucial measure to preserve airway functionality and protection during HFJV and to prevent complications [43].

To ensure safe HFJV application, anaesthetists and clinical staff require thorough technical knowledge and regular training. Proper use of ventilators, especially in critical situations and with at-risk patients, is crucial to mitigate risks and optimise the benefits of highfrequency jet ventilation [17, 21, 26].

## 5 Conclusions

This systematic review examines the safety and efficacy of high frequency jet ventilation, an unconventional ventilation technique increasingly utilised in specialised fields such as airway surgery, interventional imaging, and intensive care. Drawing on 40 studies in adult and paediatric medicine, the review evaluates HFJV both as a standalone method and in comparison, with conventional ventilation techniques.

The findings highlight significant advantages of HFJV, particularly its ability to maintain low alveolar pressures and enhance surgical conditions in procedures requiring minimal organ movement. These features make HFJV particularly suited for airway surgeries and situations where precise control of ventilation parameters and surgical access are crucial for optimal oxygenation and successful outcomes.

However, the review also emphasises the challenges and risks associated with HFJV. Accurate adjustment of ventilation parameters is critical, as inappropriate settings may lead to complications such as pneumothorax. In airway surgeries, laser-associated risks, such as airway fires causing fatal burns, must be carefully managed. The complexity of the technique and the need for specialised training among medical staff are additional barriers to its broader clinical adoption.

While HFJV demonstrates substantial benefits in specific surgical and intensive care applications, further research is necessary to define its optimal use and understand its long-term clinical implications. This could enhance its safety, efficacy, and overall reliability.

This review provides valuable insights into HFJV’s benefits and risks, offering a strong foundation for evidence-based recommendations and guiding its future clinical applications.

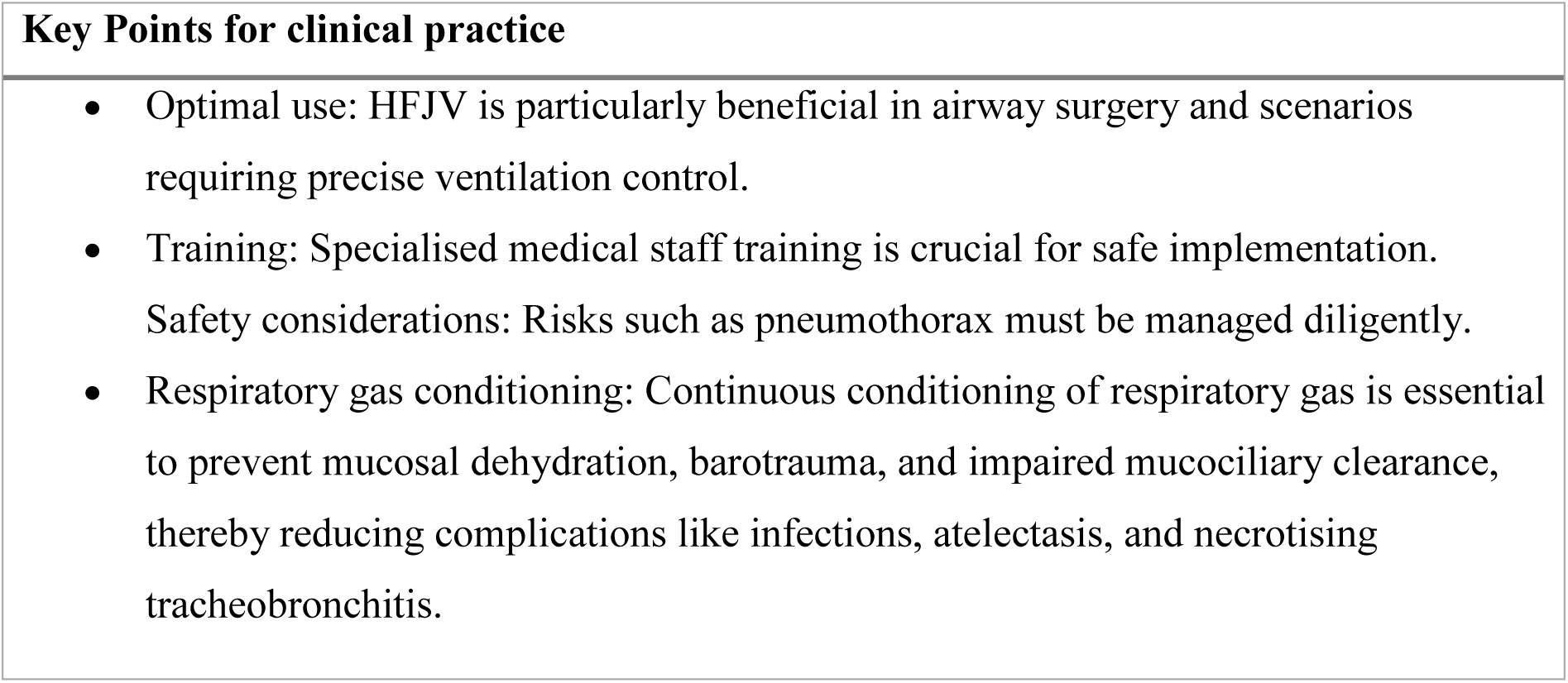

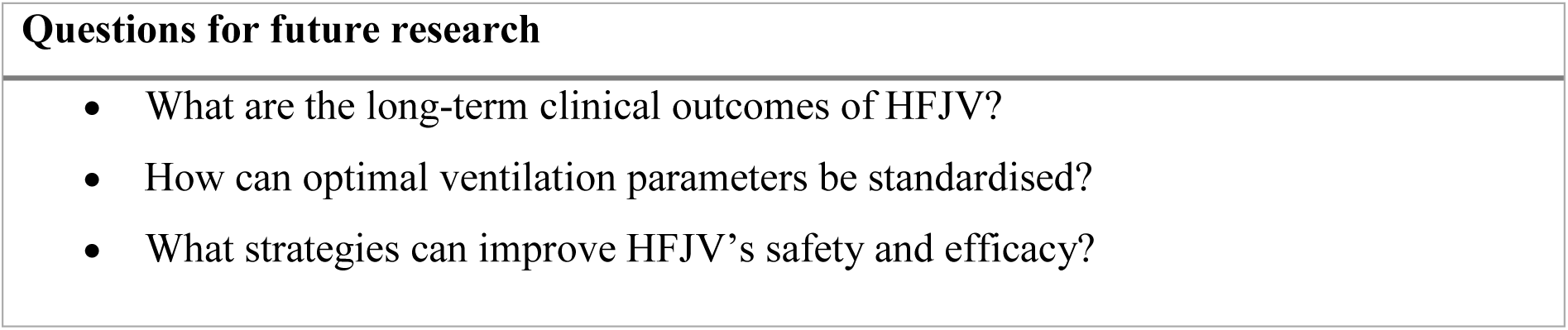

## Supporting information

Disclosure of Interest

## Data Availability

All data produced in the present work are contained in the manuscript

## Notes

### Competing Interest Statement

Competing interests: All authors have completed the ICMJE uniform disclosure form at www.icmje.org/coi_disclosure.pdf and declare: no support from any organization for the submitted work; no financial relationships with any organizations that might have an interest in the submitted work in the previous three years; no other relationships or activities that could appear to have influenced the submitted work.

### Funding Statement

This study did not receive any funding

## References

1 Evans E, Biro P, Bedforth N. Jet ventilation. Continuing Education in Anaesthesia Critical Care & Pain 2007; 7: 2–5.

2. Larsen R, Ziegenfuß T. Beatmung: Indikationen - Techniken - Krankheitsbilder [Internet]. Berlin, Heidelberg: Springer Berlin Heidelberg; 2013 [cited 2024 May 8]. Available from: https://link.springer.com/10.1007/978-3-642-29662-8.

3. AWMF online Portal der wissenschaftlichen Medizin [Internet]. AWMF Leitlinien- Register [cited 2024 May 8]. Available from: https://register.awmf.org/de/start.

4 Deutsche Gesellschaft für Hals-Nasen-Ohren-Heilkunde, Kopf-und Hals-Chirurgie. S1-Leitlinie: Tracheo-Bronchoskopie [Internet]. Oxford, UK: Oxford Centre for Evidence-Based Medicine; 2015 JulReport No.: 017/061. Available from: https://www.awmf.org/leitlinien.

5. Drifa Freysdottir, MD. High Frequency Jet Ventilator (HFJV) for Neonates Clinical Pathway. Johns Hopkins All Children’s Hospital; 2022 Dec.

6. ISO 80601-2-87:2021(en), Medical electrical equipment — Part 2-87: Particular requirements for basic safety and essential performance of high-frequency ventilators [Internet]. [cited 2024 May 8]. Available from: https://www.iso.org/obp/ui/en/#iso:std:iso:80601:-2-87:ed-1:v1:en.

7 Godet T, Jabaudon M, Blondonnet R, et al. High frequency percussive ventilation increases alveolar recruitment in early acute respiratory distress syndrome: an experimental, physiological and CT scan study. Crit Care 2018; 22: 3.

8. Murthy PR, Ak AK. High Frequency Ventilation. StatPearls [Internet] Treasure Island (FL): StatPearls Publishing; 2024 [cited 2024 May 8]. Available from: http://www.ncbi.nlm.nih.gov/books/NBK563151/.

9 Lucangelo U, Fontanesi L, Antonaglia V, et al. High frequency percussive ventilation (HFPV). Principles and technique. Minerva Anestesiol 2003; 69: 841–848, 848–851.

10 Myers M, Rodrigues N, Ari A. High-frequency oscillatory ventilation: A narrative review. CJRT 2019; 55: 40–46.

11 Musil P, Harsanyi S, Torok P, et al. Application and Technical Principles of Catheter High-Frequency Jet Ventilation. ARM 2023; 91: 278–287.

12 Miller AG, Bartle RM, Rehder KJ. High-Frequency Jet Ventilation in Neonatal and Pediatric Subjects: A Narrative Review. Respir Care 2021; 66: 845–856.

13 Page MJ, McKenzie JE, Bossuyt PM, et al. The PRISMA 2020 statement: an updated guideline for reporting systematic reviews. BMJ 2021; : n71.

14 Booth A, Sutton A, Papaioannou D. Systematic approaches to a successful literature review. Second edition. Los Angeles: Sage; 2016.

15 Atkinson LZ, Cipriani A. How to carry out a literature search for a systematic review: a practical guide. BJPsych advances 2018; 24: 74–82.

16. Oxford 2011 Levels of Evidence [Internet]. 2011. Available from: https://www.cebm.net/.

17 Bourgain JL, Desruennes E, Fischler M, et al. Transtracheal high frequency jet ventilation for endoscopic airway surgery: a multicentre study. British Journal of Anaesthesia 2001; 87: 870–875.

18 Zhang Q, Macartney J, Sampaio L, et al. High Frequency Jet Ventilation during Initial Management, Stabilization, and Transport of Newborn Infants with Congenital Diaphragmatic Hernia: A Case Series. Critical Care Research and Practice 2013; 2013: 1– 5.

19 Denys A, Lachenal Y, Duran R, et al. Use of High-Frequency Jet Ventilation for Percutaneous Tumor Ablation. Cardiovasc Intervent Radiol 2014; 37: 140–146.

20 Chung DYF, Tse DML, Boardman P, et al. High-Frequency Jet Ventilation under General Anesthesia Facilitates CT-Guided Lung Tumor Thermal Ablation Compared with Normal Respiration under Conscious Analgesic Sedation. Journal of Vascular and Interventional Radiology 2014; 25: 1463–1469.

21 Helmstaedter V, Tellkamp R, Schwab B, et al. Die Hochfrequenz-Jet-Ventilation in der HNO-Heilkunde – chirurgische und anästhesiologische Aspekte. Laryngo-Rhino-Otologie 2014; 93: 455–460.

22 Kampe S, Rocha M, Darwiche K, et al. [Sufficient high frequency jet ventilation during a period of 2.5 h - Airway management during resection of a tracheaesophageal fistula and tracheal resection]. Anasthesiologie, Intensivmedizin, Notfallmedizin, Schmerztherapie: AINS 2016; 51: 368–371.

23 Munoz-Acuna R, Tartler TM, Azizi BA, et al. Recovery and safety with prolonged high-frequency jet ventilation for catheter ablation of atrial fibrillation: A hospital registry study from a New England healthcare network. Journal of Clinical Anesthesia 2024; 93: 111324.

24. Maseri A, Delhez Q, Hardy M. Re: Recovery and safety with prolonged high-frequency jet ventilation for catheter ablation of atrial fibrillation: A hospital registry study from a New England healthcare network. Journal of Clinical Anesthesia 2024; 99: 111667.

25 Anvekar AP, Shah PS, Nathan EA, et al. High frequency jet ventilation in preterm infants: experience from Western Australia. The Journal of Maternal-Fetal & Neonatal Medicine 2019; 32: 2824–2829.

26 Wheeler CR, Smallwood CD, O’Donnell I, et al. Assessing Initial Response to High-Frequency Jet Ventilation in Premature Infants With Hypercapnic Respiratory Failure. Respiratory Care 2017; 62: 867–872.

27 Plavka R, Dokoupilová M, Pazderová L, et al. High-Frequency Jet Ventilation Improves Gas Exchange in Extremely Immature Infants with Evolving Chronic Lung Disease. American Journal of Perinatology 2006; 23: 467–472.

28 Rosal Martins M, Van Boven M, Schmitz S, et al. Technical description of a modified jet ventilation injector for airway laser surgery in neonates and infants: retrospective analysis of 20 cases. Journal of Clinical Anesthesia 2016; 32: 142–147.

29 Wheeler CR, Stephens H, O’Donnell I, et al. Mortality Risk Factors in Preterm Infants Treated with High-Frequency Jet Ventilation. Respir Care 2020; 65: 1631–1640.

30 Yang M, Wang B, Hou Q, et al. High frequency jet ventilation through mask contributes to oxygen therapy among patients undergoing bronchoscopic intervention under deep sedation. BMC Anesthesiology 2021; 21: 65.

31 El-Tahan MR, Doyle DJ, Hassieb AG. High-frequency jet ventilation using the Arndt bronchial blocker for refractory hypoxemia during one-lung ventilation in a myasthenic patient with asthma. Journal of Clinical Anesthesia 2014; 26: 570–573.

32 Fearnley RA, Badiger S, Oakley RJ, et al. Elective use of the Ventrain for upper airway obstruction during high-frequency jet ventilation. Journal of Clinical Anesthesia 2016; 33: 233–235.

33 Aizer A, Qiu JK, Cheng AV, et al. Rapid pacing and high-frequency jet ventilation additively improve catheter stability during atrial fibrillation ablation. Journal of Cardiovascular Electrophysiology 2020; 31: 1678–1686.

34. Rojas-Reyes MX, Orrego-Rojas PA. Rescue high-frequency jet ventilation versus conventional ventilation for severe pulmonary dysfunction in preterm infants. Cochrane Neonatal Group, ed. Cochrane Database of Systematic Reviews [Internet] 2015 [cited 2024 May 31]; 2015. Available from: http://doi.wiley.com/10.1002/14651858.CD000437.pub3.

35 Sivasambu B, Hakim JB, Barodka V, et al. Initiation of a High-Frequency Jet Ventilation Strategy for Catheter Ablation for Atrial Fibrillation. JACC: Clinical Electrophysiology 2018; 4: 1519–1525.

36 Gilbey P, Kukuev Y, Samet A, et al. The quality of the surgical field during functional endoscopic sinus surgery—The effect of the mode of ventilation—A randomized, prospective, double-blind study. The Laryngoscope 2009; 119: 2449–2453.

37 Volpi S, Tsoumakidou G, Loriaud A, et al. Electromagnetic navigation system combined with High-Frequency-Jet-Ventilation for CT-guided hepatic ablation of small US-Undetectable and difficult to access lesions. International Journal of Hyperthermia 2019; 36: 1050–1056.

38 Elgie LD, McPherson K, Yeung J, et al. In-circuit high-frequency jet ventilation to reduce organ motion in a child undergoing sarcoma ablation. Anaesthesia Reports 2021; 9: 55–58.

39. Altinsoy S, Sayin MM, Özkan D, et al. Is HFJV a better alternative ventilation technique for percutaneous dilatational tracheostomy? A randomized trial. Minerva Anestesiologica [Internet] 2022 [cited 2024 May 31]; 88. Available from: https://www.minervamedica.it/index2.php?show=R02Y2022N07A0588.

40 Wang H, Yang C, Zhang B, et al. Efficacy of target-controlled infusion of propofol and remifentanil with high frequency jet ventilation in fibre-optic bronchoscopy. Singapore Medical Journal 2013; 54: 689–694.

41. Conlon DCE. High Frequency Jet Ventilation: Anaesthesia Tutorial of the Week 271. Central Manchester University Hospitals, UK; 2012 Oct.

42 Aloy A, Schragl E. Jet-Ventilation [Internet]. 1995 [cited 2024 Nov 11]. Available from: http://link.springer.com/10.1007/978-3-7091-9355-6.

43 Biro’ P, Wiedemann K. Jetve tilation und Anästhesie für diagnostische und therapeutische Eingriffe an den Atemwegen. 1999; 48: 669–685.

